# HIV testing uptake according to opt-in, opt-out or risk-based testing approaches: a systematic review and meta-analysis

**DOI:** 10.1101/2022.03.10.22272235

**Authors:** Qi Rui Soh, Leon Y.J. Oh, Eric P.F. Chow, Cheryl C. Johnson, Muhammad S. Jamil, Jason J. Ong

## Abstract

**Purpose of review:** Improving HIV testing uptake is essential to ending the HIV pandemic. HIV testing approaches can be opt-in, opt-out or risk-based. This systematic review examines and compares the uptake of HIV testing in opt-in, opt-out and risk-based testing approaches.

**Recent findings:** There remains missed opportunities for HIV testing in a variety of settings using different approaches: opt-in (a person actively accepts to be tested for HIV), opt-out (a person is informed that HIV testing is routine/standard of care, and they actively decline if they do not wish to be tested for HIV) or risk-based (using risk-based screening tools to focus testing on certain individuals or sub-populations at greater risk of HIV). It is not clear how the approach could impact HIV test uptake when adjusted for other factors (e.g. rapid testing, country-income level, test setting and population tested).

**Summary:** We searched four databases for studies reporting on HIV test uptake. In total, 18,238 records were screened, and 150 studies were included in the review. Most studies described an opt-in approach (87 estimates), followed by opt-out (76) and risk-based (19). Opt-out testing was associated with 64.3% test uptake (*I*^*2*^=99.9%), opt-in testing with 59.8% (*I*^*2*^=99.9%), and risk-based testing with 54.4% (*I*^*2*^=99.9%). When adjusted for settings that offered rapid testing, country income level, setting and population tested, opt-out testing had a significantly higher uptake (+12% (95% confidence intervals: 3-21), *p*=0.007) than opt-in testing. We also found that emergency department patients and hospital outpatients had significantly lower HIV test uptake than other populations.

## INTRODUCTION

Optimizing HIV testing services is critical for ending the HIV/AIDS pandemic. Testing informs people living with HIV (PLHIV) of their status, preferably during the early stages of infection.(1) Earlier HIV detection and management has many benefits, including reducing morbidity and mortality, and preventing onward transmission.(2) It is more cost-effective to detect HIV infection early, as late presentations result in significantly higher medical costs and incur more public health expenditure.(1) Knowing one’s HIV-negative status also enables use of effective biomedical prevention strategies like pre-exposure prophylaxis.(3)

Despite the importance of HIV testing, many countries are not on track to meet the Joint United Nations Programme on HIV/AIDS 95-95-95 targets where 95% of PLHIV know their HIV status, 95% of people who know their status are receiving treatment, and 95% of people on treatment have a supressed viral load.(4) Globally, it is estimated that 84% of PLHIV were aware of their HIV status, with 87% of these were receiving treatment and 90% of these were virologically suppressed in 2020.(5) HIV/AIDS-related deaths have only declined by 57.5%, from 1.9 million in 2010 to ∼680,000 in 2020.(5) Even in well-resourced health systems, a significant proportion of PLHIV are still diagnosed late.(6) In particular, the uptake of HIV testing services remain low in key populations, resulting from structural issues that limit access and fear of stigmatisation and breach of confidentiality.(7) Discriminatory attitudes towards PLHIV persist and negatively impact the use of HIV services.(8) Further, the fear of HIV-related stigma has led to PLHIV avoiding disclosure of HIV status and delaying or staying in treatment.(9)

HIV testing services should always be voluntary and can take several approaches: opt-in (a person actively accepts to be tested for HIV), opt-out (a person is informed that HIV testing is routine/standard of care, and they actively decline if they do not wish to be tested for HIV) or risk-based (using risk-based screening tools to focus testing on certain individuals or sub-populations at greater risk of HIV).(10) Since 2006, the United States Centers for Disease Control and Prevention (CDC) has recommended an “opt-out” approach, in which voluntary HIV testing is a part of routine health care for individuals between the ages of 13 and 64.(11) Previous studies have suggested that this screening policy might reduce stigma by normalising HIV testing and making it a common behaviour.(12-14) Similarly, since 2007, the WHO recommends an opt-out approach to offer provider-initiated HIV testing service in health facilities for: (1) all patients, irrespective of epidemic setting, whose clinical presentation might result from underlying HIV infection; (2) as a standard part of medical care for all patients attending health facilities in high HIV prevalence settings; and (3) more selectively in low HIV prevalence settings.(15) Alternatively, a risk-based approach uses a set of criteria to either identify at-risk individuals for HIV testing who would not otherwise be offered a test (“screen in”) or exclude people from a routine offer of a test (“screen out”).(10)

This systematic review examined the uptake of HIV testing by comparing opt-in, opt-out or risk-based testing approaches.

## METHODS

### Search strategy and selection criteria for the systematic literature review

Ovid MEDLINE, Ovid EMBASE, Web of Science and Global Health were searched between 1^st^ January 2010 to 9^th^ July 2020. The search terminology revolved around two key aspects: “HIV” and “Risk assessments or screening”. Appendix 1 shows the full search strategy. The inclusion criteria were any study that contained primary data on the uptake of HIV testing among those offered testing; we then grouped this according to opt-in, opt-out and risk-based testing. Systematic literature reviews, editorials, duplicated results from the same study, laboratory studies about HIV diagnostic performance, and studies restricting study populations by clinical outcomes (e.g., men with urethritis or women with cervicitis) were excluded. The primary outcome of interest was the uptake of HIV testing among those offered testing.

Titles and abstracts were independently assessed for eligibility by two reviewers (QS, LO). Another reviewer (JO) resolved any discrepancies. This systematic review has been registered at the International Prospective Register of Systematic Reviews (PROSPERO: CRD42020187838).

### Data analysis

An extraction file was created in Microsoft Excel and the following information was collected: country income level, setting of the study, population tested, whether testing was opt-in/opt-out/risk-based, and presence of rapid testing. Data extraction was conducted by two reviewers (QS, LO), and another reviewer (JO) resolved any discrepancies. The quality of each study was also assessed by two reviewers (QS, LO) using the relevant critical appraisal tool from Johanna Briggs Institute.(16)

### Statistical analysis

We used descriptive analysis to summarise the characteristics of the studies included. We used the Fisher exact probability test to assess for statistically significant differences according to the testing approach. A country with a high HIV prevalence was defined as having a national prevalence above 5%, as reported by UNAIDS.(17) We used random effects meta-analysis to calculate the pooled proportion of people tested for HIV according to the type of HIV testing approach (opt-in, opt-out, risk-based). Inter-study heterogeneity was assessed using the *I*^*2*^ statistic. We explored heterogeneity using subgroup analysis and meta-regression according to availability of rapid HIV testing, country-income level, study setting, population targeted, and the latest study year. Publication bias was assessed using funnel plot and Egger’s test. STATA version 16 (StataCorp. 2019. *Stata Statistical Software: Release 16*. College Station, TX: StataCorp LLC) was used to perform all statistical analyses. This review is reported per Preferred Reporting Items for Systematic Reviews and Meta-Analyses (PRISMA) guidelines.(18)

## RESULTS

The initial search identified 18,238 potential articles; and 150 were included in this systematic review (Figure 1). Figure 2 summarizes the country of origin of the studies. Majority of studies arose from North America (n=83), followed by Africa (n=32) and Europe (n=20).

**Figure 1.**
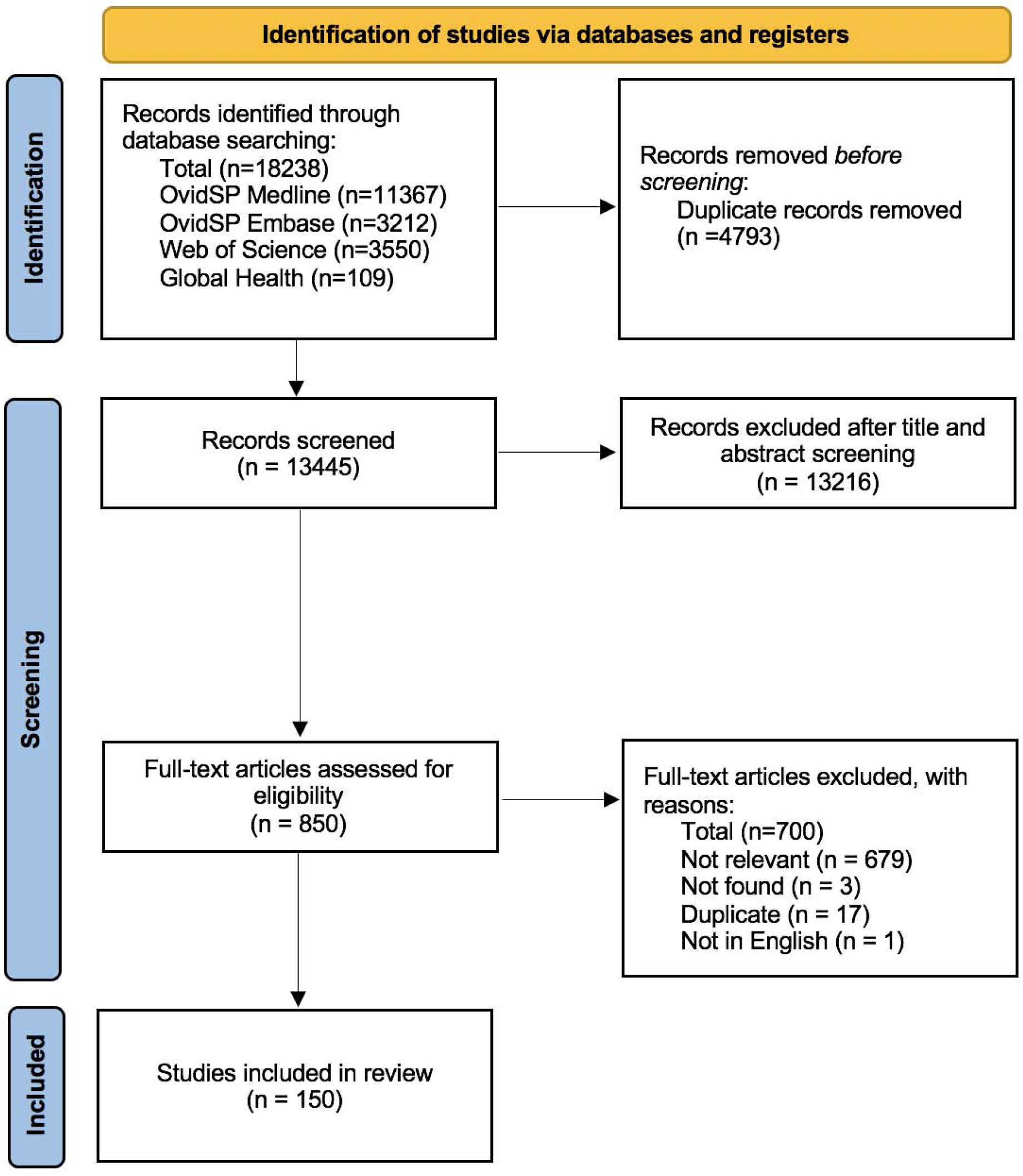
PRISMA Flow diagram

**Figure 2.**
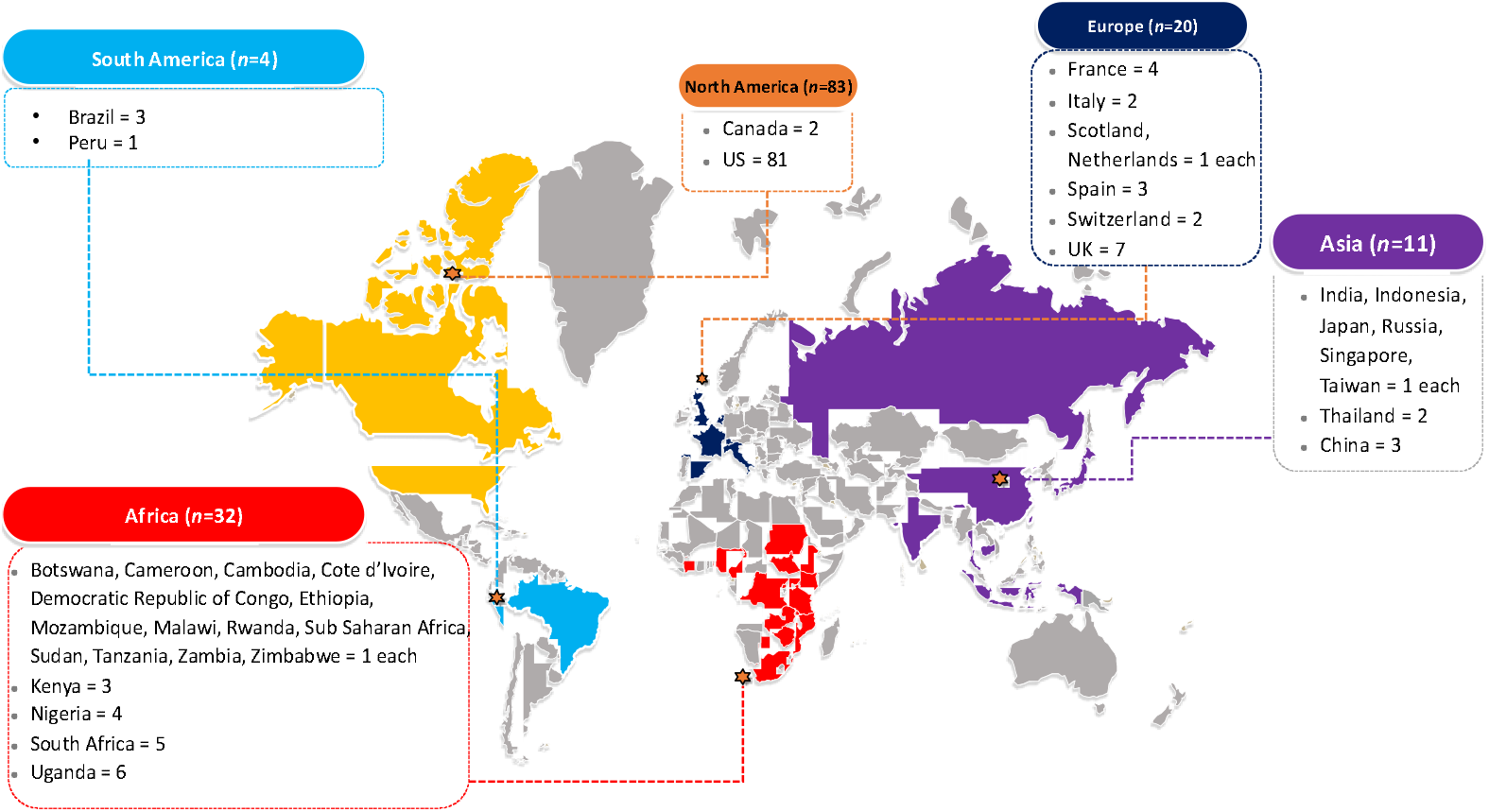
Countries of included studies (N=150)

Table 1 summarises the characteristics of the included studies according to the country’s HIV prevalence. Most studies were from high-(71%) and middle-income countries (22%), conducted in the emergency department (ED) (39%), for ED patients (41%), and involved settings with rapid testing (58%).

**Table 1.**
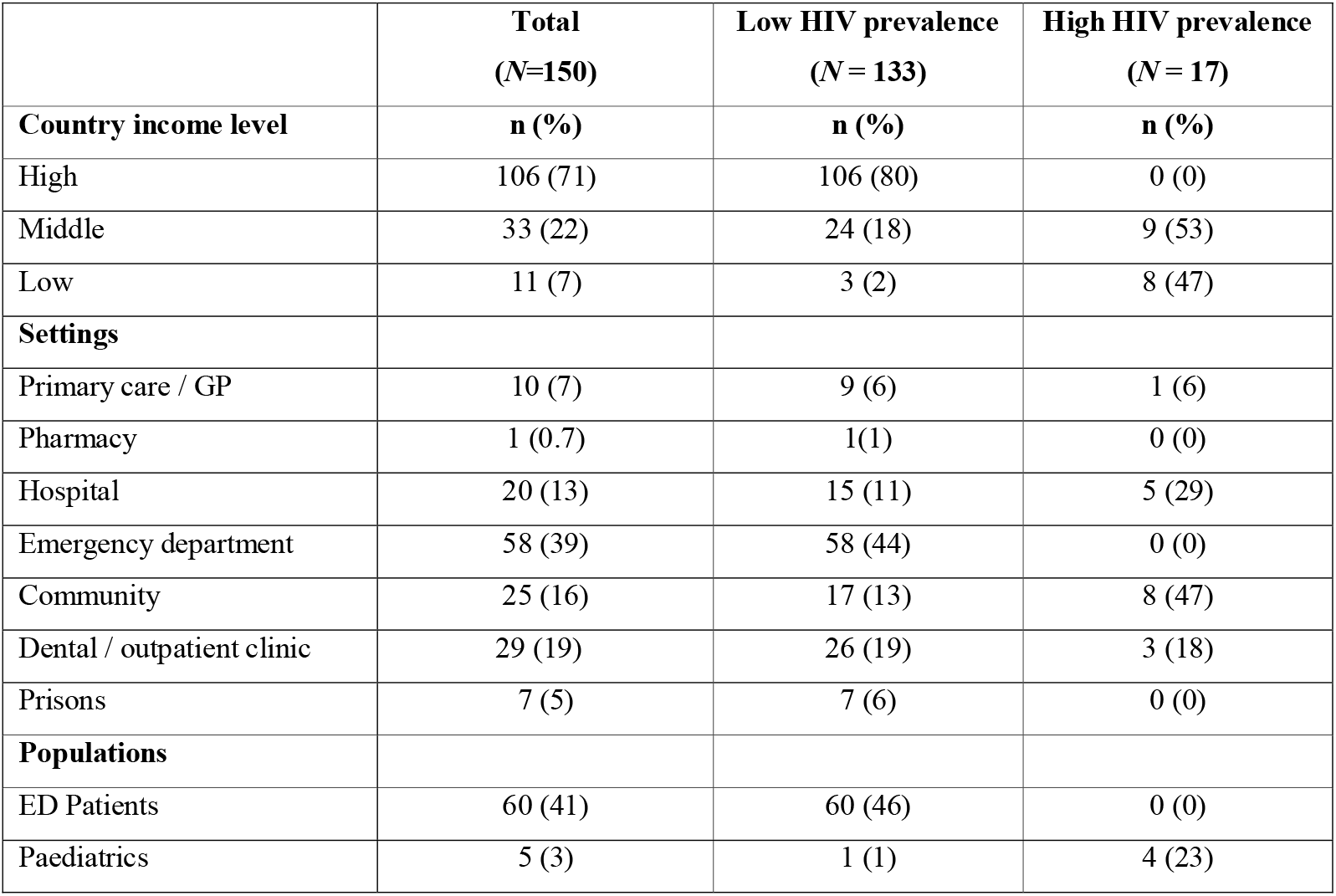
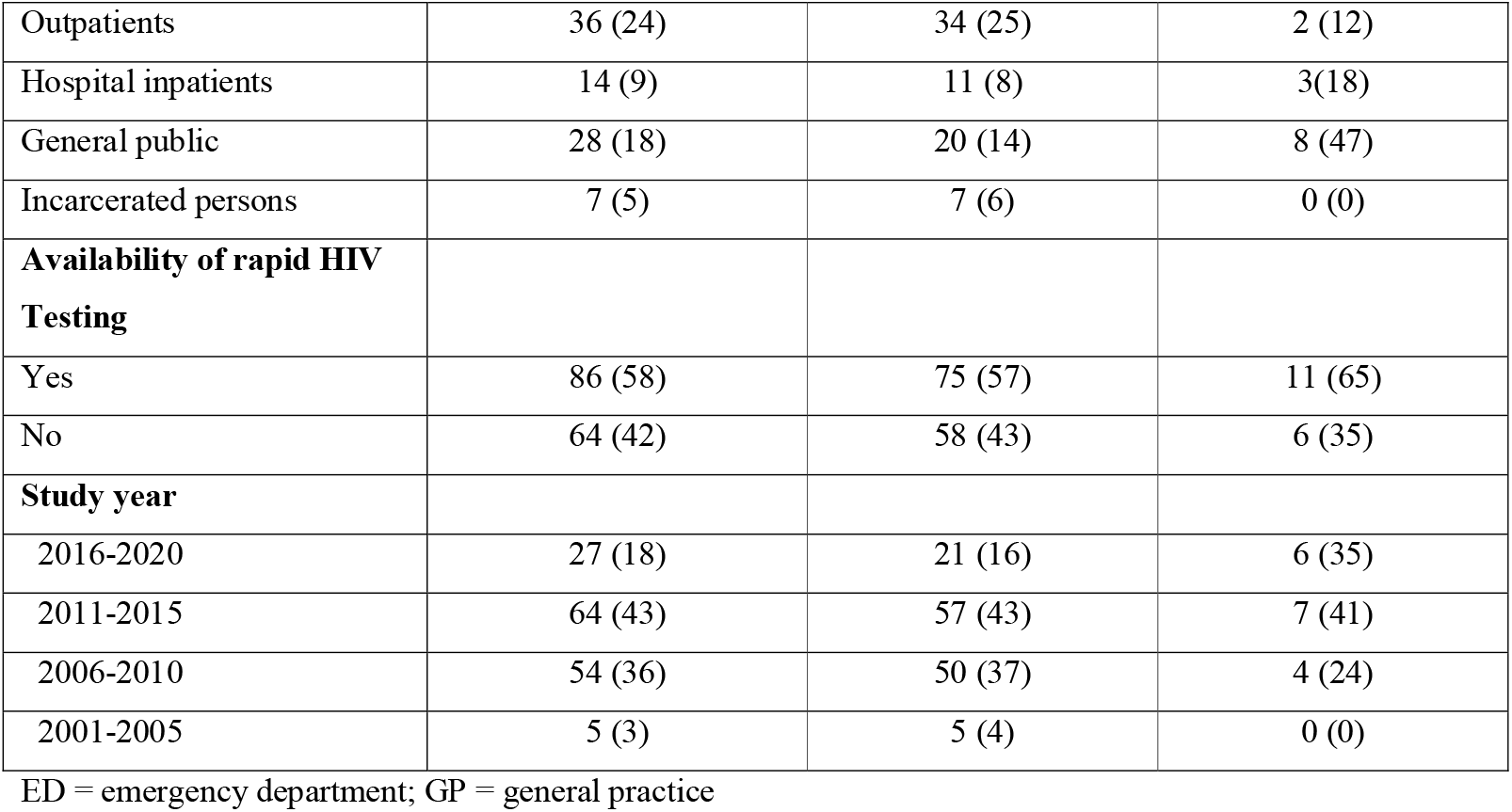
Study characteristics, according to low- and high-(≥5%) HIV prevalence(19)

Table 2 compares the study characteristics of opt-in, opt-out and risk-based testing. We found that more studies from high-income countries used opt-out or risk-based approaches, and more studies from community-based settings and those targeting the general public used the opt-in approach.

**Table 2.** Study characteristics according to opt-in testing, opt-out testing or risk-based testing approaches.

|  | **Opt-in**<br>**(N = 87)** | **Opt-out**<br>**(N = 76)** | **Risk-based**<br>**(N = 19)** | **P value** |
| --- | --- | --- | --- | --- |
| **Country income level** | n (%) | n (%) | n (%) |  |
| High | 52 (60) | 62 (82) | 15 (79) | 0.007 |
| Middle | 25 (29) | 11 (14) | 4 (21) | 0.083 |
| Low | 10 (11) | 3 (4) | 0 (0) | 0.104 |
| **Settings** |  |  |  |  |
| Primary care / GP | 4 (5) | 6 (8) | 4 (21) | 0.050 |
| Pharmacy | 1 (1) | 0 (0) | 0 (0) | 1.000 |
| Hospital | 7 (8) | 12 (16) | 4 (21) | 0.131 |
| Emergency department | 29 (33) | 34 (45) | 7 (38) | 0.311 |
| Community | 23 (26) | 5 (6) | 2 (10) | 0.002 |
| Dental / outpatient clinic | 18 (20) | 12 (16) | 1 (5) | 0.281 |
| Prisons | 5 (7) | 7 (9) | 1 (5) | 0.769 |
| **Populations** |  |  |  |  |
| ED patients | 30 (34) | 33 (44) | 6 (32) | 0.432 |
| Paediatrics | 1 (1) | 3 (4) | 2 (10) | 0.090 |
| Outpatients | 18 (20) | 19 (25) | 7 (38) | 0.309 |
| Hospital inpatients | 5 (6) | 10 (13) | 1 (5) | 0.218 |
| General public | 28 (32) | 4 (5) | 2 (10) | <0.001 |
| Incarcerated persons | 5 (7) | 7 (9) | 1 (5) | 0.769 |
| **Rapid HIV testing** |  |  |  |  |
| Yes | 57 (65) | 39 (52) | 9 (47) | 0.135 |
| No | 30 (35) | 37 (48) | 10 (53) | - |

Note: of 150 unique studies, some evaluated more than one approach and thus will appear more than once in the columns.

Table 3 summarises the pooled proportion of people testing for HIV according to various settings. It demonstrates that opt-out testing had higher uptake of people testing for HIV compared with opt-in and risk-based testing (64.3% vs. 59.8%), although it was not statistically significantly different. However, in the meta-regression analysis (Table 4), when we adjusted for rapid HIV testing, country income level, test setting, population tested and the year of study, opt-out testing had a significantly higher HIV test uptake compared with opt-in and risk-based testing (additional 12% and 15%, respectively).

**Table 3.** Pooled proportion of people testing for HIV.

| | Number of studies | Pooled proportion of people testing for HIV (%) | 95% confidence interval | $I^2$ (p value) |
| --- | --- | --- | --- | --- |
| Total (N=182) |  | 61.2 | 57.4-64.9 | 99.9 (<0.001) |
| **Type of HIV testing service** |  |  |  |  |
| Opt-in | 87 | 59.8 | 52.2-67.3 | 99.9 (<0.001) |
| Opt-out | 76 | 64.3 | 57.4-70.9 | 99.9 (<0.001) |
| Risk-based | 19 | 54.4 | 41.2-67.4 | 99.9 (<0.001) |
| **Rapid testing** |  |  |  |  |
| Available | 86 | 62.2 | 56.1-68.0 | 100.0 (<0.001) |
| Not available | 64 | 60.0 | 54.4-65.5 | 100.0 (<0.001) |
| **Country income level** |  |  |  |  |
| High | 106 | 53.7 | 49.5-57.8 | 100.0 (<0.001) |
| Middle | 33 | 80.4 | 73.9-86.2 | 99.9 (<0.001) |
| Low | 11 | 69.1 | 57.2-79.9 | 99.6 (<0.001) |
| **Setting** |  |  |  |  |
| Hospital | 22 | 72.0 | 56.4-85.2 | 100.0 (<0.001) |
| GP/primary care | 14 | 81.4 | 72.0-89.3 | 98.5 (<0.001) |
| Pharmacy | 1 | 39.5 | 33.2-46.1 | - |
| Community-based | 30 | 79.2 | 73.6-84.3 | 100.0 (<0.001) |
| Emergency Department | 71 | 46.6 | 40.1-53.3 | 100.0 (<0.001) |
| Prison | 14 | 68.8 | 49.9-84.9 | 100.0 (<0.001) |
| Outpatients | 29 | 55.9 | 44.9-66.7 | 100.0 (<0.001) |
| Mixed | 3 | 46.9 | 16.1-79.1 | - |
| **Populations** |  |  |  |  |
| Inpatients | 15 | 68.4 | 45.7-87.3 | 100.0 (<0.001) |
| Emergency patients | 69 | 47.2 | 40.6-53.9 | 100.0 (<0.001) |
| Paediatrics | 6 | 75.7 | 63.1-86.4 | 99.7 (<0.001) |
| General public | 34 | 76.4 | 71.1-81.3 | 100.0 (<0.001) |
| Outpatients | 41 | 64.8 | 50.1-78.2 | 100.0 (<0.001) |
| Incarcerated persons | 11 | 68.8 | 49.9-84.9 | 100.0 (<0.001) |
| Mixed | 3 | 46.9 | 16.1-79.1 | - |
| **Latest study year** |  |  |  |  |
| 2016-2020 | 27 | 69.2 | 57.8-79.5 | 100.0 (<0.001) |
| 2011-2015 | 64 | 59.8 | 54.7-64.8 | 100.0 (<0.001) |
| 2006-2010 | 54 | 58.2 | 48.7-67.3 | 100.0 (<0.001) |
| 2001-2005 | 5 | 63.2 | 49.7-75.7 | 99.8 (<0.001) |

**Supplementary Table 1.** Meta-regression of HIV test uptake.

| Variable | Univariable |  |  | Multivariable<sup>1</sup> |  |
| --- | --- | --- | --- | --- | --- |
| | $\beta$ (95% CI) | P-value | Adjusted R<sup>2</sup> | $\beta$ (95% CI) | P-value |
| **Type of HIV testing service approach** |  |  | -0.15% |  |  |
| Opt-in | Reference |  |  | Reference |  |
| Opt-out | 0.04 (-0.05 to 0.12) | 0.413 |  | 0.12 (0.03 to 0.21) | 0.007 |
| Risk-based | -0.05 (-0.19 to 0.09) | 0.506 |  | -0.03 (-0.17 to 0.11) | 0.685 |
| **Rapid testing** |  |  | -0.51% |  |  |
| Available | Reference |  |  | Reference |  |
| Not available | -0.02 (-0.10 to 0.07) | 0.674 |  | -0.03 (-0.11 to 0.06) | 0.542 |
| **Country income level** |  |  | 12.1% |  |  |
| High | Reference |  |  | Reference |  |
| Middle | 0.24 (0.15 to 0.33) | <0.001 |  | 0.21 (0.09 to 0.32) | 0.001 |
| Low | 0.14 (-0.01 to 0.30) | 0.067 |  | 0.19 (0.00 to 0.38) | 0.051 |
| Mixed | 0.28 (-0.23 to 0.79) | 0.273 |  | 0.17 (-0.41 to 0.75) | 0.560 |
| **Setting** |  |  | 17.9% |  |  |
| Hospital | Reference |  |  | Reference |  |
| GP/primary care | 0.10 (-0.07 to 0.28) | 0.251 |  | 0.04 (-0.22 to 0.29) | 0.779 |
| Pharmacy | -0.30 (-0.82 to 0.22) | 0.256 |  | -0.20 (-0.76 to 0.36) | 0.487 |
| Community-based | 0.06 (-0.08 to 0.21) | 0.371 |  | 0.03 (-0.22 to 0.29) | 0.795 |
| Emergency Department | -0.23 (-0.35 to -0.10) | <0.001 |  | -0.35 (-0.68 to -0.03) | 0.032 |
| Prison | -0.03 (-0.20 to 0.14) | 0.761 |  | 0.09 (-0.11 to 0.28) | 0.373 |
| Outpatients | -0.15 (-0.29 to 0.0) | 0.044 |  | -0.29 (-0.54 to -0.04) | 0.021 |
| Mixed | -0.23 (-0.54 to 0.08) | 0.141 |  | -0.32 (-0.67 to 0.03) | 0.072 |
| **Populations** |  |  | 11.5% |  |  |
| Inpatients | Reference |  |  | Reference |  |
| Emergency patients | -0.18 (-0.33 to -0.03) | 0.017 |  | 0.26 (-0.07 to 0.59) | 0.124 |
| Paediatrics | 0.09 (-0.16 to 0.34) | 0.495 |  | -0.01 (-0.18 to 0.38) | 0.958 |
| General public | 0.07 (-0.09 to 0.23) | 0.383 |  | 0.10 (-0.18 to 0.38) | 0.487 |
| Outpatients | -0.03 (-0.19 to 0.13) | 0.710 |  | 0.22 (-0.05 to 0.48) | 0.108 |
| Incarcerated persons | 0.01 (-0.18 to 0.21) | 0.900 |  | * |  |
| Mixed | -0.19 (-0.52 to 0.14) | 0.251 |  | * |  |
| **Latest study year** |  |  | -0.16% |  |  |
| 2016-2020 | Reference |  |  | Reference |  |
| 2011-2015 | -0.07 (-0.19 to 0.04) | 0.194 |  | 0.01 (-0.10 to 0.12) | 0.818 |
| 2006-2010 | -0.09 (-0.20 to 0.02) | 0.114 |  | 0.04 (-0.08 to 0.15) | 0.552 |
| 2001-2005 | -0.03 (-0.29 to 0.23) | 0.812 |  | 0.19 (-0.06 to 0.44) | 0.135 |

\* omitted because of collinearity
<sup>1</sup> Adjusted $R^2$ 23.3%

Supplementary Figure 1 shows the funnel plot which demonstrates a possibility for publication bias with under-reporting of studies with lower HIV test uptake. The quality assessment for each paper is presented in Supplementary Tables 1-3.

## DISCUSSION

This systematic review aimed to understand the uptake of HIV testing by comparing opt-in, opt-out and risk-based testing approaches. This study adds to the evidence base regarding HIV testing approaches. We found that opt-out testing (when adjusted for rapid testing, country income level, setting and population tested) had higher uptake than opt-in and risk-based testing. We also found that the population of emergency department patients and hospital outpatients had significantly lower HIV test uptake than other populations.

Our finding that opt-out testing for HIV was associated with a higher proportion of people testing than opt-in testing is consistent with other studies. For example, a 2017 systematic review and meta-analysis comparing HIV opt-out testing and opt-in testing amongst patients attending emergency departments found that the opt-out strategies had higher uptake (44%) than the opt-in strategies (19%).(20) We extend the evidence base for the value of opt-out testing, as we included studies from various settings beyond emergency departments. The value of opt-out testing is exemplified by a 2021 study in Kenya that reported a 2.2-fold greater odds of new HIV diagnosis using opt-out point of care than opt-in testing.(21) The study reported higher refusal rates for opt-in testing, while a higher proportion of participants in the opt-out testing were willing to disclose risky sexual practices, suggesting that they were more likely to participate if testing were presented as part of standard care.(21) The study also reported that physicians were more likely to offer tests to patients who are at a higher risk of HIV (i.e. never tested, tested >1 year ago, older men), and therefore were likely to miss a substantial proportion during opt-in testing.(21)

Our review found that opt-out testing was mostly implemented in the emergency department setting. Yet, HIV test uptake was the lowest in emergency departments compared with other settings where opt-out testing was available. Whilst there could be value in HIV testing in emergency departments, studies have shown HIV testing in emergency departments could have poor linkage to care,(22) low test acceptance rates among marginalised populations,(23) high cost per positive diagnosis,(24) and lack of cultural competency being integrated.(25) This could also be due to the transient nature of conditions and acute care needed in the emergency department, where the focus is on the patient’s current issue and less on peripheral issues like HIV testing. Furthermore, HIV testing uptake could be higher when a physician offers the test (26-28) which may not always be the case in a busy emergency department. Nevertheless, an ED-based HIV screening program remains an integral component of the overall HIV screening strategy to reduce the current HIV testing gap and complement existing community-based HIV screening programs. Therefore, our study highlights the need for further improvements for HIV testing beyond opt-out testing strategies for the emergency department setting.

These are missed opportunities for HIV testing in certain settings. Our review uncovered that pharmacies, followed by primary care clinics, had the lowest uptake of HIV testing. In many countries, the majority of the population sees a primary care practitioner at least once a year.(29, 30) The literature surrounding insights into GPs’ current HIV testing practices reveals the barriers GPs face to routinely offering testing, including being worried about potentially harming patient relationships(31) and feeling incapable of offering HIV tests due to perceived poor knowledge.(32) Steps should be taken to address barriers around HIV testing in primary care to improve HIV detection rates.(33, 34) In addition, pharmacies can provide point-of-care HIV testing(35) and participate in HIV prevention related to pre-exposure prophylaxis and post-exposure prophylaxis.(36)

One unexpected finding from our review was in settings where rapid testing was available, there was no significant difference in HIV test uptake compared to settings without rapid testing. This observation should be interpreted with caution. One possibility could be because the majority of studies with rapid testing were conducted in emergency department settings, a setting with the lowest testing uptake in this review. Another possibility is that unlike other studies which specifically assessed the impact of rapid testing compared with venepuncture, our systematic review examined the difference in HIV test uptake between settings where rapid testing was available compared with settings that did not have rapid testing uptake. Therefore, there could be other confounders related to subpopulations attending these settings.(37, 38) There is evidence of greater appeal of rapid testing compared with venepuncture. For example, a 2013 systematic review on rapid point-of-care HIV testing found that youth preferred rapid point-of-care tests compared to traditional testing methods.(39) Similarly, a study of adults attending general practices in France reported higher acceptability of a rapid test (92%) compared with venepuncture (64%).(40) Studies report that patients prefer to receive their results quickly and would recommend rapid testing to their peers.(41, 42) Rapid testing can reach high-risk populations in clinical and community settings, which is critical in testing untested individuals.(43, 44) However, there is evidence that some patients may have concerns regarding the reliability of the rapid test and having their clinical visits prolonged.(33) Further research is warranted to understand how rapid testing (including HIV self-testing) could improve HIV testing rates using an opt-out approach.

There are a few limitations of this systematic review. First, many studies included were from high-income countries, specifically, more than half were from the United States. As such, the results may not be easily generalisable to other settings and/or in lower-income settings. A large proportion (71 of 150) of studies were from an emergency department. Therefore, our findings could be skewed by the large proportion of studies from the United States, emergency department settings and/or high-income countries. Second, we found a low number of studies using the risk-based HIV testing approach (19 of 150 articles), thus exposing a gap in the literature for future studies to evaluate the value of this approach.(10) Third, we found high heterogeneity between studies, highlighting the importance of the need for local, contextualised evidence when deciding between an opt-in, opt-out or risk-based testing approach. We explored this heterogeneity in our meta-regression analyses and found that country-income level, settings and type of population could explain some of this variability, but there remain unexplained confounders.

In conclusion, this review adds to the current literature that opt-out testing can significantly improve HIV test uptake compared to opt-in in various settings and across different populations. We also uncovered settings (emergency department, primary care, pharmacy) where HIV test uptake remains poor, highlighting the need to implement new strategies in those settings to improve HIV test uptake if we are to end the HIV/AIDS pandemic.

## Data Availability

All data produced in the present study are available upon reasonable request to the authors

## DECLARATIONS

### Availability of data

All relevant data are presented in the manuscript and online supplementary materials. Any further details can be obtained by contacting the corresponding author.

### Code availability

Not applicable

### Authors’ contributions

JJO, CJ, MSJ conceptualized the idea. LO and QS performed the screening and extraction of data. JJO analysed the data. LO and QS wrote the original draft. All authors contributed to the writing of the manuscript and approved the final version for submission.

### Ethics approval

Not applicable

### Consent to participate

Not applicable

### Consent for publication

Not applicable

** Of major importance

## REFERENCES

1. Apers H, Nöstlinger C, Van Beckhoven D, Deblonde J, Apers L, Verheyen K, et al. Identifying key elements to inform HIV-testing interventions for primary care in Belgium. Health promotion international. 2020;35(2):301–11.

2. Brault MA, Spiegelman D, Hargreaves J, Nash D, Vermund SH. Treatment as Prevention: Concepts and Challenges for Reducing HIV Incidence. J Acquir Immune Defic Syndr. 2019;82 Suppl 2:S104–S12.

3. Chou R, Evans C, Hoverman A, Sun C, Dana T, Bougatsos C, et al. Preexposure Prophylaxis for the Prevention of HIV Infection: Evidence Report and Systematic Review for the US Preventive Services Task Force. JAMA. 2019;321(22):2214–30.

4. UNAIDS. Understanding Fast-Track. Accelerating action to end the AIDS epidemic by 2030 [Available from: https://www.unaids.org/sites/default/files/media_asset/201506_JC2743_Understanding_FastTrack_en.pdf.

5. HIV/AIDS JUNPo. Fact sheet: World AIDS Day 2019—global HIV statistics. Dec 1, 2019. 2020.

6. Late Presentation Working Groups in Euro S, Cohere. Estimating the burden of HIV late presentation and its attributable morbidity and mortality across Europe 2010-2016. BMC Infect Dis. 2020;20(1):728.

7. Krause J, Subklew-Sehume F, Kenyon C, Colebunders R. Acceptability of HIV self-testing: a systematic literature review. BMC public health. 2013;13(1):1–9.

8. Chesney MA, Smith AW. Critical delays in HIV testing and care: The potential role of stigma. American behavioral scientist. 1999;42(7):1162–74.

9. Tee Y, Huang M. Knowledge of HIV/AIDS and attitudes towards people living with HIV among the general staff of a public university in Malaysia. SAHARA-J: Journal of Social Aspects of HIV/AIDS. 2009;6(4):179–87.

10. Ong JJ, Coulthard K, Quinn C, Tang MJ, Huynh T, Jamil MS, et al. Risk-Based Screening Tools to Optimise HIV Testing Services: a Systematic Review. Curr HIV/AIDS Rep. 2022. This systematic review summarizes the accuracy of risk-based screening tools for HIV testing.

11. Prevention CfDCa. HIV Testing 2021 [Available from: https://www.cdc.gov/hiv/testing/index.html.

12. Copenhaver MM, Fisher JD. Experts outline ways to decrease the decade-long yearly rate of 40,000 new HIV infections in the US. AIDS and Behavior. 2006;10(1):105–14.

13. Hutchinson AB, Corbie-Smith G, Thomas SB, Mohanan S, Del Rio C. Understanding the patient’s perspective on rapid and routine HIV testing in an inner-city urgent care center. AIDS Education and prevention. 2004;16(2):101–14.

14. Irwin KL, Valdiserri RO, Holmberg SD. The acceptability of voluntary HIV antibody testing in the United States: a decade of lessons learned. Aids. 1996.

15. UNAIDS, WHO HIV/AIDS Programme. Guidance on Provider-Initiated Testing and Counseling in Health Facilities. World Health Organization; May, 2007. [Available from: http://apps.who.int/iris/bitstream/handle/10665/43688/9789241595568_eng.pdf?sequence=1.

16. Johanna Briggs Institute Critical Appraisal Tools [Available from: https://jbi.global/critical-appraisal-tools.

17. UNAIDS. Global HIV & AIDS statistics - fact sheet. [Available from: https://www.unaids.org/en/resources/fact-sheet#:~:text=In%202020%2C%20there%20were%2037.7,HIV%20were%20women%20and%20girls.

18. Page MJ, McKenzie JE, Bossuyt PM, Boutron I, Hoffmann TC, Mulrow CD, et al. The PRISMA 2020 statement: An updated guideline for reporting systematic reviews. PLoS Med. 2021;18(3):e1003583.

19. UNAIDS. Global HIV & AIDS statistics-2020 fact sheet 2020 [

20. Henriquez-Camacho C, Villafuerte-Gutierrez P, Perez-Molina JA, Losa J, Gotuzzo E, Cheyne N. Opt-out screening strategy for HIV infection among patients attending emergency departments: systematic review and meta-analysis. HIV Med. 2017;18(6):419–29. This systematic review compares opt-out versus opt-in screening in the emergency department.

21. Sanders EJ, Agutu C, van der Elst E, Hassan A, Gichuru E, Mugo P, et al. Effect of an opt□out point□of□care HIV□1 nucleic acid testing intervention to detect acute and prevalent HIV infection in symptomatic adult outpatients and reduce HIV transmission in Kenya: a randomized controlled trial. HIV medicine. 2021.

22. Menon AA, Nganga□Good C, Martis M, Wicken C, Lobner K, Rothman RE, et al. Linkage□to□care Methods and Rates in US Emergency Department–based HIV Testing Programs: A Systematic Literature Review Brief Report. Academic Emergency Medicine. 2016;23(7):835–42.

23. Merchant RC, Seage III GR, Mayer KH, Clark MA, DeGruttola VG, Becker BM. Emergency department patient acceptance of opt-in, universal, rapid HIV screening. Public Health Reports. 2008;123(3_suppl):27–40.

24. Cowan E, Herman HS, Rahman S, Zahn J, Leider J, Calderon Y. Bundled HIV and hepatitis C testing in the emergency department: a randomized controlled trial. Western Journal of Emergency Medicine. 2018;19(6):1049.

25. Merchant RC, Liu T, Clark MA, Carey MP. Facilitating HIV/AIDS and HIV testing literacy for emergency department patients: a randomized, controlled, trial. BMC emergency medicine. 2018;18(1):1–18.

26. White DA, Scribner AN, Vahidnia F, Dideum PJ, Gordon DM, Frazee BW, et al. HIV screening in an urban emergency department: comparison of screening using an opt-in versus an opt-out approach. Ann Emerg Med. 2011;58(1 Suppl 1):S89–95.

27. Simpson WM, Johnstone FD, Boyd FM, Goldberg DJ, Hart GJ, Prescott RJ. Uptake and acceptability of antenatal HIV testing: randomised controlled trial of different methods of offering the test. Bmj. 1998;316(7127):262–7.

28. Haukoos JS, Hopkins E, Eliopoulos VT, Byyny RL, LaPerriere KA, Mendoza MX, et al. Development and Implementation of a Model to Improve Identification of Patients Infected with HIV Using Diagnostic Rapid Testing in the Emergency Department. Academic Emergency Medicine. 2007;14(12):1149–57.

29. Statistics ABo. Patient Experiences in Australia: Summary of Findings 2020 [cited 2021 20/08/2021]. Available from: https://www.abs.gov.au/statistics/health/health-services/patient-experiences-australia-summary-findings/latest-release.

30. Irving G, Neves AL, Dambha-Miller H, Oishi A, Tagashira H, Verho A, et al. International variations in primary care physician consultation time: a systematic review of 67 countries. BMJ Open. 2017;7(10):e017902.

31. Hindocha S, Charlton T, Rayment M, Theobald N. Feasibility and acceptability of routine human immunodeficiency virus testing in general practice: your views. Primary health care research & development. 2013;14(2):212–6.

32. Manirankunda L, Loos J, Debackaere P, Nöstlinger C. “It is not easy”: challenges for provider-initiated HIV testing and counseling in Flanders, Belgium. AIDS Education and Prevention. 2012;24(5):456–68.

33. Avery AK, Del Toro M, Caron A. Increases in HIV screening in primary care clinics through an electronic reminder: an interrupted time series. BMJ quality & safety. 2014;23(3):250–6.

34. Glew S, Pollard A, Hughes L, Llewellyn C. Public attitudes towards opt-out testing for HIV in primary care: a qualitative study. British Journal of General Practice. 2014;64(619):e60–e6.

35. McKeirnan K, Kherghehpoush S, Gladchuk A, Patterson S. Addressing Barriers to HIV Point-of-Care Testing in Community Pharmacies. Pharmacy (Basel). 2021;9(2).

36. Myers JE, Farhat D, Guzman A, Arya V. Pharmacists in HIV Prevention: An Untapped Potential. Am J Public Health. 2019;109(6):859–61.

37. Sharma M, Ong JJ, Celum C, Terris-Prestholt F. Heterogeneity in individual preferences for HIV testing: A systematic literature review of discrete choice experiments. EClinicalMedicine. 2020;29-30:100653.

38. Ong JJ, Nwaozuru U, Obiezu-Umeh C, Airhihenbuwa C, Xian H, Terris-Prestholt F, et al. Designing HIV Testing and Self-Testing Services for Young People in Nigeria: A Discrete Choice Experiment. Patient. 2021;14(6):815–26.

39. Turner SD, Anderson K, Slater M, Quigley L, Dyck M, Guiang CB. Rapid point-of-care HIV testing in youth: a systematic review. J Adolesc Health. 2013;53(6):683–91.

40. Demorat H, Lopes A, Chopin D, Delcey V, Clevenbergh P, Simoneau G, et al. Acceptability and feasibility of HIV testing in general medicine by ELISA or rapid test from finger-stick whole blood. Presse Med. 2018;47(2):e15–e23.

41. Merchant RC, Clark MA, Seage III GR, Mayer KH, Degruttola VG, Becker BM. Emergency department patient perceptions and preferences on opt-in rapid HIV screening program components. AIDS care. 2009;21(4):490–500.

42. Smith LV, Rudy ET, Javanbakht M, Uniyal A, Sy LS, Horton T, et al. Client satisfaction with rapid HIV testing: comparison between an urban sexually transmitted disease clinic and a community-based testing center. AIDS Patient Care & STDs. 2006;20(10):693–700.

43. Mutch AJ, Lui C-W, Dean J, Mao L, Lemoire J, Debattista J, et al. Increasing HIV testing among hard-to-reach groups: examination of RAPID, a community-based testing service in Queensland, Australia. BMC health services research. 2017;17(1):1–7.

44. Pottie K, Medu O, Welch V, Dahal GP, Tyndall M, Rader T, et al. Effect of rapid HIV testing on HIV incidence and services in populations at high risk for HIV exposure: an equity-focused systematic review. BMJ open. 2014;4(12):e006859.

