## Supplementary file for "HIV testing uptake according to opt-in, opt-out or risk-based testing approaches: a systematic review and meta-analysis"

**APPENDIX 1**

**Medline Ovid – 9 July 2020**

1. (HIV or "human immunodeficiency virus" or "acquired immunodeficiency" or "acquired immune deficiency" or AIDS or "hiv infect*" or "acquired immunodeficiency syndrome" or "acquired immunedeficiency syndrome" or "acquired immuno-deficiency syndrome" or "acquired immune-deficiency syndrome" or "HIV-infected" or "HIV-positive" or "HIV/AIDS" or "HIV-1" or "HIV-2" or hiv1 or hiv2 or "HIV diagnos*" or serodiagnos* or "HIV acquisition" or "acquir* HIV").ti,ab.

2. *HIV Infections/di or *Acquired Immunodeficiency Syndrome/di or *HIV Seropositivity/di or *AIDS Serodiagnosis/ or *HIV Infections/pc

3. 1 or 2

4. (screen* or test or tests or tested or testing).ti,ab.

5. *Mass Screening/mt or *Diagnostic Tests, Routine/mt

6. (Risk* or high-risk).ti,ab.

7. *Risk Assessment/ or *Risk Factors/ or *Risk-taking/ or *Risk/

8. 4 or 5 or 6 or 7

9. (tool* or assess* or scoring or score or validation or predict* or profil* or stratification or decision* or "artificial intelligence" or algorithm* or calculat* or detect* or comput* or question* or scale* or evaluat* or target* or strateg* or performance or sensitivity or specificity).ti.

10. *Decision Support Techniques/ or *"Sensitivity and Specificity"/

11. 9 or 10

12. 3 and 8 and 11

13. Animals/

14. 12 not 13

15. limit 14 to yr="2010 - 2020"

11367 results

**EMBASE Ovid – 9 July 2020**

1. (HIV or "human immunodeficiency virus" or "acquired immunodeficiency" or "acquired immune deficiency" or AIDS or "hiv infect*" or "acquired immunodeficiency syndrome" or "acquired immunedeficiency syndrome" or "acquired immuno-deficiency syndrome" or "acquired immune-deficiency syndrome" or "HIV-infected" or "HIV-positive" or "HIV/AIDS" or "HIV-1" or "HIV-2" or hiv1 or hiv2 or "HIV diagnos*" or serodiagnos* or "HIV acquisition" or "acquir* HIV").ti.

2. (screen* or test or tests or tested or testing or risk* or high-risk).ti.

3. (tool* or assess* or scoring or score or validation or predict* or profil* or stratification or decision* or artificial intelligence or algorithm* or calculat* or detect* or comput* or question* or scale* or evaluat* or target* or strateg* or performance or sensitivity or specificity).ti.

4. 1 and 2 and 3

5. animal/

6. 4 not 5

7. limit 6 to yr="2010 - 2020"

3212 results

**Web of Science – 1 July**

Indexes=SCI-EXPANDED, SSCI, A&HCI, CPCI-S, CPCI-SSH, BKCI-S, BKCI-SSH, ESCI, CCR-EXPANDED, IC Timespan=2010-2020

1.	ti=(HIV or "human immunodeficiency virus" or "acquired immunodeficiency" or "acquired immune deficiency" or AIDS or "hiv infect*" or "acquired immunodeficiency syndrome" or "acquired immunedeficiency syndrome" or "acquired immuno-deficiency syndrome" or "acquired immune-deficiency syndrome" or "HIV-infected" or "HIV-positive" or "HIV/AIDS" or "HIV-1" or "HIV-2" or hiv1 or hiv2 or "HIV diagnos*" or serodiagnos* or "HIV acquisition" or "acquir* HIV")

2.	ti=(screen* or test or tests or tested or testing or risk* or high-risk)

3.	ti=(tool* or assess* or scoring or score or validation or predict* or profil* or stratification or decision* or artificial intelligence or algorithm* or calculat* or detect* or comput* or question* or scale* or evaluat* or target* or strateg* or performance or sensitivity or specificity)

4.	#3 AND #2 AND #1

3550 results

**Global Health search – 2 July 2020**

title:(HIV or "human immunodeficiency virus" or "acquired immunodeficiency" or "acquired immune deficiency" or AIDS or "hiv infect*" or "acquired immunodeficiency syndrome" or "acquired immunedeficiency syndrome" or "acquired immuno-deficiency syndrome" or "acquired immune-deficiency syndrome" or "HIV-infected" or "HIV-positive" or "HIV/AIDS" or "HIV-1" or "HIV-2" or hiv1 or hiv2 or "HIV diagnos*" or serodiagnos* or "HIV acquisition" or "acquir* HIV") AND title:(screen* or test or tests or tested or testing or risk* or high-risk) AND title:(tool* or assess* or scoring or score or validation or predict* or profil* or stratification or decision* or artificial intelligence or algorithm* or calculat* or detect* or comput* or question* or scale* or evaluat* or target* or strateg* or performance or sensitivity or specificity) AND yr:[2010 TO 2020]

109 results

**Supplementary Figure 1 Funnel plot**

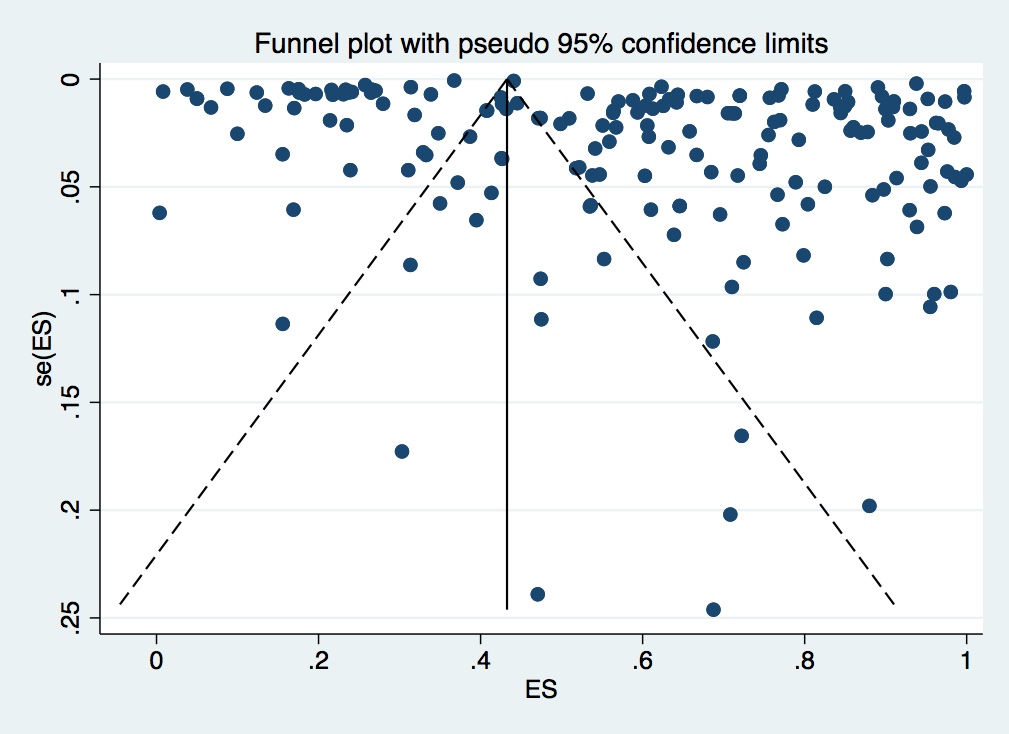

Egger’s test = 0.020

**Supplementary Table 1 Quality assessment for cross-sectional studies.**

| Paper | 1 | 2 | 3 | 4 | 5 | 6 | 7 | 8 |
| --- | --- | --- | --- | --- | --- | --- | --- | --- |
| Abramson (1) | Y | Y | Y | Y | NA | NA | Y | Y |
| Ahmed (2) | Y | Y | Y | Y | Y | Y | Y | Y |
| Allan-Blitz (3) | Y | Y | Y | Y | Y | Y | Y | Y |
| Amesty (4) | Y | Y | Y | Y | Y | Y | Y | Y |
| Anim (5) | Y | Y | Y | Y | U | U | Y | Y |
| Aparicio (6) | Y | Y | Y | Y | Y | Y | Y | Y |
| Aronson (7) | Y | Y | Y | Y | Y | Y | Y | Y |
| Ayieko (8) | Y | Y | Y | Y | Y | U | Y | Y |
| Babatunde (9) | Y | Y | Y | Y | Y | Y | Y | Y |
| Baisley (10) | Y | Y | Y | Y | Y | U | Y | Y |
| Batey (11) | Y | Y | Y | Y | Y | U | Y | Y |
| Beckwith (12) | Y | Y | Y | Y | Y | U | Y | Y |
| Bender (13) | Y | Y | Y | Y | Y | Y | Y | Y |
| Blackstock (14) | Y | Y | Y | Y | Y | U | Y | Y |
| Boyer (15) | Y | U | Y | Y | Y | U | Y | Y |
| Boyer (16) | Y | Y | U | U | U | U | Y | Y |
| Brigstock-Barron (17) | Y | Y | Y | Y | U | Y | Y | Y |
| Brondani (18) | Y | Y | Y | Y | U | Y | Y | Y |
| Brown (19) | Y | Y | Y | U | U | Y | Y | Y |
| Bryce (20) | Y | Y | Y | Y | Y | Y | Y | Y |
| Buzi (21) | Y | Y | Y | Y | Y | Y | Y | Y |
| Casalino (22) | Y | Y | U | NA | NA | NA | Y | Y |
| Cayuelas (23) | Y | Y | U | Y | Y | Y | Y | Y |
| CDC (24) | Y | Y | U | Y | Y | U | Y | Y |
| CDC (25) | Y | Y | Y | U | Y | U | Y | Y |
| Chhagan (26) | Y | Y | Y | Y | U | U | Y | Y |
| Cochand (27) | Y | Y | Y | Y | Y | Y | Y | Y |
| Collaboration (28) | Y | Y | U | Y | Y | U | Y | Y |
| Coppola (29) | Y | Y | U | Y | U | U | Y | Y |
| Costello (30) | Y | Y | Y | Y | Y | U | Y | Y |
| Cowan (31) | Y | Y | Y | Y | U | U | Y | Y |
| Crumby (32) | Y | Y | Y | Y | Y | U | Y | Y |
| Cunningham (33) | Y | Y | Y | Y | Y | Y | Y | Y |
| D'Almeida (34) | Y | Y | Y | Y | Y | Y | Y | Y |
| Dalal (35) | Y | Y | Y | Y | Y | Y | Y | Y |
| Ezeanolue (36) | Y | Y | Y | Y | Y | U | Y | Y |
| Felsen (37) | Y | Y | Y | Y | Y | Y | Y | Y |
| Gbadamosi (38) | Y | Y | Y | Y | Y | Y | Y | Y |
| Geoffroy (39) | Y | Y | Y | Y | Y | Y | Y | Y |
| Geren (40) | U | U | Y | U | U | U | Y | Y |
| Geren (41) | Y | Y | Y | Y | Y | Y | Y | Y |
| Gilbert (42) | Y | Y | Y | Y | U | U | Y | Y |
| Gwadz (43) | Y | Y | Y | Y | Y | U | Y | Y |
| Harmon (44) | Y | Y | Y | Y | Y | Y | Y | Y |
| Hector (45) | Y | Y | Y | Y | Y | U | Y | Y |
| Hempling (46) | Y | Y | Y | Y | U | U | Y | Y |
| Hsieh (47) | Y | Y | Y | Y | U | U | Y | Y |
| Hsieh (48) | Y | Y | Y | Y | U | U | Y | Y |
| Hurley (49) | Y | Y | Y | Y | U | U | Y | Y |
| Ibrahim (50) | Y | Y | Y | Y | U | U | Y | Y |
| Idris (51) | Y | Y | Y | Y | Y | Y | Y | Y |
| Kabami (28) | Y | Y | Y | Y | Y | U | Y | Y |
| Kancheya (52) | Y | Y | Y | Y | Y | U | Y | Y |
| Kayigamba (53) | Y | Y | Y | Y | Y | Y | Y | Y |
| Kennedy (54) | Y | Y | Y | Y | Y | U | Y | Y |
| Khawcharoenporn (55) | Y | Y | Y | Y | Y | Y | Y | Y |
| Kinsler (56) | Y | Y | Y | Y | Y | Y | Y | Y |
| Kurth (57) | Y | Y | Y | Y | Y | U | Y | Y |
| Lander (58) | Y | Y | Y | Y | U | U | Y | Y |
| Leblanc (59) | Y | Y | Y | Y | Y | Y | Y | Y |
| Leonard (60) | Y | Y | Y | Y | U | Y | Y | Y |
| Levin (61) | Y | Y | Y | Y | U | U | Y | Y |
| Lolekha (62) | Y | Y | Y | Y | Y | U | Y | Y |
| Lubelchek (63) | Y | Y | Y | Y | Y | Y | Y | Y |
| Lucas (64) | Y | Y | Y | Y | Y | Y | Y | Y |
| Luiken (65) | Y | Y | Y | Y | Y | U | Y | Y |
| Lyons (66) | Y | Y | Y | Y | Y | U | Y | Y |
| Lyss (67) | Y | Y | Y | Y | Y | Y | Y | Y |
| Malaju (68) | Y | Y | Y | Y | Y | Y | Y | Y |
| Manjezi (69) | Y | Y | Y | Y | Y | U | Y | Y |
| Mehta (70) | Y | Y | Y | Y | Y | Y | Y | Y |
| Menacho (71) | Y | Y | Y | Y | U | U | Y | Y |
| Montoy (72) | Y | Y | Y | Y | U | U | Y | Y |
| Mwembo-Tambwe (73) | Y | Y | Y | Y | U | U | Y | Y |
| Myers (74) | Y | Y | Y | Y | Y | Y | Y | Y |
| Nakigudde (75) | Y | Y | Y | Y | U | U | Y | Y |
| Naugle (76) | Y | Y | Y | Y | N | N | Y | Y |
| Nelwan (77) | U | Y | Y | Y | N | N | Y | Y |
| Nsirim (78) | N | Y | Y | Y | N | N | Y | Y |
| Nunn (79) | Y | Y | Y | Y | N | N | Y | Y |
| Ogbo (80) | Y | Y | Y | Y | N | N | Y | Y |
| Osorio (81) | Y | Y | Y | Y | N | N | Y | Y |
| Palfreeman (82) | N | Y | Y | Y | N | N | Y | Y |
| Pant Pai (83) | Y | Y | Y | Y | N | N | Y | Y |
| Pant Pai (84) | Y | Y | Y | Y | N | N | Y | Y |
| Pontiff (85) | N | N | U | U | N | N | Y | Y |
| Prekker (86) | Y | Y | Y | Y | N | N | Y | Y |
| Prekker (87) | N | N | U | U | N | N | Y | Y |
| Raman (88) | N | N | U | U | N | N | Y | Y |
| Ribeiro (89) | Y | Y | Y | Y | N | N | Y | Y |
| Rodriguez (90) | N | Y | Y | Y | N | N | Y | Y |
| Rosen (91) | Y | Y | Y | Y | Y | Y | Y | Y |
| Russell (92) | N | N | U | U | N | N | Y | Y |
| Russo (93) | Y | Y | Y | Y | Y | U | Y | Y |
| Sankoff (94) | Y | Y | U | U | N | N | Y | Y |
| Sattin (95) | Y | Y | Y | Y | N | N | Y | Y |
| Sau (96) | Y | Y | Y | Y | U | U | Y | Y |
| Schlesinger (97) | Y | Y | U | U | N | N | Y | Y |
| Schnall (98) | U | Y | Y | Y | N | N | Y | Y |
| Shih (99) | Y | Y | Y | Y | N | N | Y | Y |
| Silva (100) | Y | Y | Y | Y | N | N | Y | Y |
| Spaulding (101) | N | Y | Y | Y | N | N | Y | U |
| Strick (25) | N | Y | Y | Y | N | N | Y | U |
| Takano (102) | Y | Y | Y | Y | N | N | Y | Y |
| Tan (103) | Y | Y | Y | U | N | N | Y | Y |
| Tlamsa (104) | Y | Y | U | U | N | N | U | U |
| Truong (105) | Y | Y | U | U | N | N | Y | Y |
| Valenti (106) | U | Y | Y | Y | N | N | Y | Y |
| Veloso (107) | Y | Y | Y | Y | N | N | Y | Y |
| Vohra (108) | N | Y | U | U | N | N | Y | U |
| Walensky (109) | Y | N | U | U | N | N | U | U |
| Wang (110) | Y | Y | Y | Y | Y | Y | Y | Y |
| Wang (111) | Y | Y | Y | Y | N | N | Y | Y |
| Wheatley (112) | Y | Y | Y | Y | N | N | Y | Y |
| White (113) | Y | Y | Y | Y | N | N | Y | Y |
| White (114) | Y | Y | Y | Y | N | N | Y | Y |
| White (115) | Y | Y | Y | Y | N | N | Y | Y |
| White (116) | Y | Y | Y | Y | N | N | Y | Y |
| Wilbur (117) | Y | Y | Y | Y | N | N | Y | Y |
| Xia (118) | U | Y | Y | Y | N | N | Y | Y |
| Yumo (119) | Y | Y | Y | Y | N | N | Y | Y |
| Zhu (120) | Y | Y | Y | U | N | N | Y | Y |
| Zimet (121) | U | Y | Y | U | N | N | Y | U |
| Zimmerman (122) | Y | Y | y | U | N | N | Y | U |

Legend for table:

1. Were the criteria for inclusion in the sample clearly defined?
2. Were the study subjects and the setting described in detail?
3. Was the exposure measured in a valid and reliable way?
4. Were objective, standard criteria used for measurement of the condition?
5. Were confounding factors identified?
6. Were strategies to deal with confounding factors stated?
7. Were the outcomes measured in a valid and reliable way?
8. Was appropriate statistical analysis used?

Y – Yes

N – No

U – Unclear

NA – Not applicable

**Supplementary Table 2 Quality assessment for cohort studies**

| **Paper** | **1** | **2** | **3** | **4** | **5** | **6** | **7** | **8** | **9** | **10** | **11** |
| --- | --- | --- | --- | --- | --- | --- | --- | --- | --- | --- | --- |
| Berhie (123) | Y | Y | Y | U | U | Y | Y | U | U | Y | Y |
| CDC (25) | Y | Y | U | N | N | Y | U | U | U | U | U |
| Dominguez-Berjon (124) | Y | Y | Y | N | N | Y | Y | Y | Y | N | Y |
| Felsen (37) | Y | Y | Y | U | U | Y | Y | NA | NA | NA | Y |
| Hankin (125) | Y | U | U | U | U | U | U | U | U | U | U |
| Haukoos (126) | Y | Y | Y | N | N | Y | Y | Y | Y | N | Y |
| Haukoos (127) | Y | Y | Y | NA | NA | Y | Y | Y | Y | N | Y |
| Haukoos (128) | Y | Y | Y | N | N | Y | Y | Y | N | N | Y |
| Haukoos (129) | Y | Y | Y | N | N | Y | Y | Y | N | N | Y |
| Hechter (130) | Y | Y | Y | U | U | Y | Y | Y | Y | N | Y |
| Hoxhaj (131) | Y | Y | Y | N | N | Y | Y | Y | Y | N | Y |
| Mahajan (132) | Y | Y | Y | N | N | Y | Y | Y | N | N | Y |
| Mbopi-Keou (133) | Y | Y | U | N | N | Y | U | NA | NA | NA | U |
| McGuire (134) | NA | NA | NA | N | N | NA | NA | NA | NA | NA | Y |
| Schrantz (135) | Y | Y | Y | N | N | Y | Y | Y | Y | N | Y |
| Yeganeh (136) | NA | NA | NA | N | N | Y | Y | Y | N | Y | Y |

Legend for table:

1. Were the two groups similar and recruited from the same population?
2. Were the exposures measured similarly to assign people to both exposed and unexposed groups?
3. Was the exposure measured in a valid and reliable way?
4. Were confounding factors identified?
5. Were strategies to deal with confounding factors stated?
6. Were the groups/participants free of the outcome at the start of the study (or at the moment of exposure)?
7. Were the outcomes measured in a valid and reliable way?
8. Was the follow up time reported and sufficient to be long enough for outcomes to occur?
9. Was follow up complete, and if not, were the reasons to loss to follow up described and explored?
10. Were strategies to address incomplete follow up utilized?
11. Was appropriate statistical analysis used?

Y – Yes

N – No

NA – Not applicable

U - Unclear

**Supplementary Table 3 Quality assessment for randomized controlled trials**

| **Paper** | **1** | **2** | **3** | **4** | **5** | **6** | **7** | **8** | **9** | **10** | **11** | **12** | **13** |
| --- | --- | --- | --- | --- | --- | --- | --- | --- | --- | --- | --- | --- | --- |
| Chaffin (137) | U | U | U | Y | N | N | U | U | U | U | U | U | Y |
| Chamie (138) | Y | Y | Y | Y | N | N | Y | Y | N | Y | Y | Y | Y |
| Chamie (139) | U | U | U | U | U | U | U | U | U | U | Y | U | U |
| Chesang (140) | U | U | U | U | N | N | U | U | U | U | Y | U | U |
| Cowan (141) | Y | Y | Y | Y | Y | N | Y | Y | U | Y | Y | Y | Y |
| Gillet (142) | Y | Y | Y | Y | NA | N | Y | Y | NA | Y | Y | Y | Y |
| Haukoos (143) | U | U | U | Y | N | N | Y | Y | Y | Y | Y | U | Y |
| Kelvin (144) | U | Y | Y | Y | N | N | Y | U | U | Y | Y | Y | Y |
| Lyons (145) | Y | Y | Y | Y | N | N | Y | Y | NA | Y | Y | Y | Y |
| Merchant (146) | Y | Y | Y | Y | N | N | Y | Y | NA | Y | Y | Y | Y |

Legend for table:

1. Was true randomization used for assignment of participants to treatment groups?
2. Was allocation to treatment groups concealed?
3. Were treatment groups similar at the baseline?
4. Were participants blind to treatment assignment?
5. Were those delivering treatment blind to treatment assignment?
6. Were outcomes assessors blind to treatment assignment?
7. Were treatment groups treated identically other than the intervention of interest?
8. Was follow up complete and if not, were differences between groups in terms of their follow up adequately described and analyzed?
9. Were participants analyzed in the groups to which they were randomized?
10. Were outcomes measured in the same way for treatment groups?
11. Were outcomes measured in a reliable way?
12. Was appropriate statistical analysis used?
13. Was the trial design appropriate, and any deviations from the standard RCT design (individual randomization, parallel groups) accounted for in the conduct and analysis of the trial?

Y = Yes

N = No

U = Unclear

NA = Not applicable

1.	Abramson A, MacHtinger E. Evaluation of HIV screening utility and practicability in an inpatient medicine ward setting. Journal of Hospital Medicine. 2011;6(4 SUPPL. 2):S1.

2.	Ahmed A, Fattah S, McCallum A, Wood C, Kane S, Fleck A, et al. HIV testing in acute medicine; assessing the rates and barriers to testing in a busy Scottish acute medical unit: P140. Hiv Medicine. 2016;17.

3.	Allan-Blitz L-T, Herrera MC, Calvo GM, Vargas SK, Caceres CF, Klausner JD, et al. Venue-based HIV-testing: an effective screening strategy for high-risk populations in Lima, Peru. AIDS and Behavior. 2019;23(4):813-9.

4.	Amesty S, Crawford ND, Nandi V, Perez-Figueroa R, Rivera A, Sutton M, et al. Evaluation of pharmacy-based HIV testing in a high-risk New York City community. AIDS patient care and STDs. 2015;29(8):437-44.

5.	Anim M, Markert RJ, Okoye NE, Sabbagh W. HIV screening of patients presenting for routine medical care in a primary care setting. Journal of primary care & community health. 2013;4(1):28-30.

6.	Aparicio C, Mourez T, Simoneau G, Magnier J-D, Galichon B, Plaisance P, et al. Proposal of HIV, HBV and HCV targeted screening: short period feasibility study in a free-access outpatient medical structure. Presse medicale (Paris, France: 1983). 2012;41(10):e517-23.

7.	Aronson I, Cleland C, Rajan S, Marsch L, Bania T. Computer-based substance use reporting and acceptance of HIV testing among emergency department patients. AIDS and Behavior. 2020;24(2):475-83.

8.	Ayieko J, Chamie G, Balzer L, Kwarisiima D, Kabami J, Sang N, et al. Mobile, population-wide, hybrid HIV testing strategy increases number of children tested in rural Kenya and Uganda. The Pediatric infectious disease journal. 2018;37(12):1279.

9.	BABATUNDE OT, BABATUNDE LS, OYEDEJI OA, OWA JA. Acceptance of Human Immunodeficiency Virus Testing among Caregivers of Children using Provider-Initiated Testing and Counselling Strategy in Ido-ekiti, Nigeria: A Cross-sectional Study. Journal of Clinical & Diagnostic Research. 2019;13(8).

10.	Baisley K, Doyle AM, Changalucha J, Maganja K, Watson-Jones D, Hayes R, et al. Uptake of voluntary counselling and testing among young people participating in an HIV prevention trial: comparison of opt-out and opt-in strategies. 2012.

11.	Batey DS, Hogan VL, Cantor R, Hamlin CM, Ross-Davis K, Nevin C, et al. Short communication routine HIV testing in the emergency department: assessment of patient perceptions. AIDS research and human retroviruses. 2012;28(4):352-6.

12.	Beckwith CG, Bazerman L, Cornwall AH, Patry E, Poshkus M, Fu J, et al. An evaluation of a routine opt-out rapid HIV testing program in a Rhode Island jail. AIDS education and prevention. 2011;23(3_supplement):96-109.

13.	Ignacio RAB, Chu J, Power MC, Douaiher J, Lane JD, Collins JP, et al. Influence of providers and nurses on completion of non-targeted HIV screening in an urgent care setting. AIDS research and therapy. 2014;11(1):1-11.

14.	Blackstock OJ, King JR, Mason RD, Lee CC, Mannheimer SB. Evaluation of a rapid HIV testing initiative in an urban, hospital-based dental clinic. AIDS patient care and STDS. 2010;24(12):781-5.

15.	Boyer CB, Robles-Schrader GM, Li SX, Miller RL, Korelitz J, Price GN, et al. A comparison of network-based strategies for screening at-risk Hispanic/Latino adolescents and young adults for undiagnosed asymptomatic HIV infection. Journal of adolescent health. 2014;55(6):765-73.

16.	Boyer CB, Hightow-Weidman L, Bethel J, Li SX, Henry-Reid L, Futterman D, et al. An assessment of the feasibility and acceptability of a friendship-based social network recruitment strategy to screen at-risk African American and Hispanic/Latina young women for HIV infection. JAMA pediatrics. 2013;167(3):289-96.

17.	Brigstock-Barron O, Logan L, Sowerbutts H, Womack V, Osman M, Anderson J, et al., editors. Using epidemiology and collaborative funding to enable innovation in opportunistic screening to reduce the late diagnosis of HIV: interim results from a targeted primary care project in England (UK). JOURNAL OF THE INTERNATIONAL AIDS SOCIETY; 2016: JOHN WILEY & SONS LTD THE ATRIUM, SOUTHERN GATE, CHICHESTER PO19 8SQ, W ….

18.	Brondani M, Chang S, Donnelly L. Assessing patients’ attitudes to opt-out HIV rapid screening in community dental clinics: a cross-sectional Canadian experience. BMC research notes. 2016;9(1):1-9.

19.	Brown J, Shesser R, Simon G, Bahn M, Czarnogorski M, Kuo I, et al. Routine HIV screening in the emergency department using the new US Centers for Disease Control and Prevention Guidelines: results from a high-prevalence area. JAIDS Journal of Acquired Immune Deficiency Syndromes. 2007;46(4):395-401.

20.	Bryce G, Wilkinson P, Nicholson S, Jeffery A, Hankins M, Jackson D. A study to assess the acceptability, feasibility and cost-effectiveness of universal HIV testing with newly registering patients (aged 16-59) in primary care. HIV Med. 2011;12:3-4.

21.	Buzi RS, Madanay FL, Smith PB. Integrating routine HIV testing into family planning clinics that treat adolescents and young adults. Public Health Reports. 2016;131(1_suppl):130-8.

22.	Casalino E, Bernot B, Bouchaud O, Alloui C, Choquet C, Bouvet E, et al. Twelve months of routine HIV screening in 6 emergency departments in the Paris area: results from the ANRS URDEP study. 2012.

23.	Redondo LC, Ruíz M, Kostov B, Sequeira E, Noguera P, Herrero MA, et al. Indicator condition-guided HIV testing with an electronic prompt in primary healthcare: a before and after evaluation of an intervention. Sexually transmitted infections. 2019;95(4):238-43.

24.	Control CfD, Prevention. Rapid HIV testing in emergency departments--three US sites, January 2005-March 2006. MMWR Morbidity and mortality weekly report. 2007;56(24):597-601.

25.	Strick L, MacGowan R, Margolis A, Belcher L. HIV screening of male inmates during prison intake medical evaluation—Washington, 2006-2010. Morb Mortal Wkly Rep. 2011;60:811-3.

26.	Chhagan MK, Kauchali S, Arpadi SM, Craib MH, Bah F, Stein Z, et al. Failure to test children of HIV‐infected mothers in South Africa: implications for HIV testing strategies for preschool children. Tropical Medicine & International Health. 2011;16(12):1490-4.

27.	Cochand L, Masserey E, Bodenmann P, Troillet N. Assessment of voluntary HIV screening for asylum seekers in two Swiss cantons. Swiss medical weekly. 2019;149(0708).

28.	Kabami J, Chamie G, Kwarisiima D, Biira E, Ssebutinde P, Petersen M, et al. Evaluating the feasibility and uptake of a community‐led HIV testing and multi‐disease health campaign in rural Uganda. Journal of the International AIDS Society. 2017;20(1):21514.

29.	Coppola N, Alessio L, Gualdieri L, Pisaturo M, Sagnelli C, Caprio N, et al. A STRATEGY TO FAVOR THE ACCESS OF IRREGULAR AND REFUGEE MIGRANTS TO A SCREENING PROGRAM FOR HBV, HCV AND HIV INFECTION. Digestive and Liver Disease. 2015;47:e61-e2.

30.	Costello J, Carpentier M, Sliney A, MacLeod C, Young K, Flanigan T. Evaluation of a nurse-initiated routine HIV testing pilot on a medical-surgical unit. Medsurg Nursing. 2016;25(1):36.

31.	Cowan E, Leider J, Verma R, Perera T, Caban R, Rhee J, et al. Assessing the Effect of Being Offered Voluntary HIV Testing at the Nurse Triage Station in an Urban Hospital Emergency Department: 551. Academic Emergency Medicine. 2015;22.

32.	Crumby NS, Arrezola E, Brown EH, Brazzeal A, Sanchez TH. Experiences implementing a routine HIV screening program in two federally qualified health centers in the southern United States. Public Health Reports. 2016;131(1_suppl):21-9.

33.	Cunningham CO, Doran B, DeLuca J, Dyksterhouse R, Asgary R, Sacajiu G. Routine opt-out HIV testing in an urban community health center. AIDS Patient Care and STDs. 2009;23(8):619-23.

34.	d’Almeida KW, Kierzek G, de Truchis P, Le Vu S, Pateron D, Renaud B, et al. Modest public health impact of nontargeted human immunodeficiency virus screening in 29 emergency departments. Archives of internal medicine. 2012;172(1):12-20.

35.	Dalal S, Lee C-w, Farirai T, Schilsky A, Goldman T, Moore J, et al. Provider-initiated HIV testing and counseling: increased uptake in two public community health centers in South Africa and implications for scale-up. PloS one. 2011;6(11):e27293.

36.	Ezeanolue E, Gande N, Ekeh O. Routine HIV screening program in an urban outpatient setting. AIDS Read. 2010.

37.	Felsen UR, Torian LV, Futterman DC, Stafford S, Xia Q, Allan D, et al. An expanded HIV screening strategy in the Emergency Department fails to identify most patients with undiagnosed infection: insights from a blinded serosurvey. AIDS care. 2019.

38.	Gbadamosi SO, Itanyi IU, Menson WNA, Olawepo JO, Bruno T, Ogidi AG, et al. Targeted HIV testing for male partners of HIV-positive pregnant women in a high prevalence setting in Nigeria. PloS one. 2019;14(1):e0211022.

39.	Geoffroy E, Schell E, Jere J, Khozomba N. Going door-to-door to reach men and young people with HIV testing services to achieve the 90–90–90 treatment targets. Public health action. 2017;7(2):95-9.

40.	Geren KI, Lovecchio F, Knight J, Fromm R, Moore E, Tomlinson C, et al. Identification of acute HIV infection using fourth-generation testing in an opt-out emergency department screening program. Annals of emergency medicine. 2014;64(5):537-46.

41.	Geren KI, Moore EO, Fromm RE, Hobohm D, Jenkins J, Jordan HL, et al. Algorithms and acute infections: Innovations in routine HIV screening. Topics in Antiviral Medicine. 2014;22(E-1):304-5.

42.	Gilbert M, Alvarez M, Krajden M, Buxton J, Lester R, Money D, et al. P3. 197 Uptake and Case Detection of Prenatal Screening of Maternal Syphilis, HIV and Hepatitis C, in British Columbia, Canada, 2007–2011. Sexually Transmitted Infections. 2013;89(Suppl 1):A209-A10.

43.	Gwadz M, Cleland CM, Perlman DC, Hagan H, Jenness SM, Leonard NR, et al. Public health benefit of peer-referral strategies for detecting undiagnosed HIV infection among high-risk heterosexuals in New York City. Journal of acquired immune deficiency syndromes (1999). 2017;74(5):499.

44.	Harmon JL, Collins-Ogle M, Bartlett JA, Thompson J, Barroso J. Integrating routine HIV screening into a primary care setting in rural North Carolina. Journal of the Association of Nurses in AIDS Care. 2014;25(1):70-82.

45.	Hector J, Davies MA, Dekker-Boersema J, Aly MM, Abdala CA, Langa EBR, et al. Acceptability and performance of directly assisted HIV self-testing in adolescents in rural Mozambique. Tropical Medicine & International Health. 2017;22:368-.

46.	Hempling MC, Pakianathan M, Majewska W, Shields K, Davey S, Karim J. Pilot project evaluating hiv testing in ST george's emergency department. Emergency Medicine Journal. 2011;28(SUPPL. 1):A5.

47.	Hsieh Y, Gauvey-Kern M, Woodfield A. Novel Approach to Streamlining HIV Testing in the Emergency Department: Touch-Screen Kiosk Systems for Offering HIV Test and Risk Assessment. Academic Emergency Medicine. 2012;19:S294.

48.	Hsieh Y-H. Evaluation of a Rapid HIV Screening Program in an Urban Academic Adult Emergency Department to Identify Individuals with Undiagnosed HIV Infection.

49.	Hurley H, Gawenus L, Gardner EM, Rowan S, editors. Relationship of Risk Screening to HIV and Viral Hepatitis Detection for Participants in a Colorado Narcotic Replacement Therapy Program. Open Forum Infectious Diseases; 2017: Oxford University Press US.

50.	Ibrahim M, Maswabi K, Ajibola G, Moyo S, Hughes MD, Batlang O, et al. Targeted HIV testing at birth supported by low and predictable mother‐to‐child transmission risk in Botswana. Journal of the International AIDS Society. 2018;21(5):e25111.

51.	Idris A, Elsamani E, Elnasri A. Sociodemographic predictors of acceptance of voluntary HIV testing among pregnant women in a large maternity hospital, Omdurman, Sudan. EMHJ-Eastern Mediterranean Health Journal. 2015;21(4):273-9.

52.	Kancheya NG, Jordan AK, Zulu IS, Chanda D, Vermund SH. Improved HIV testing coverage after scale-up of antiretroviral therapy programs in urban Zambia: evidence from serial hospital surveillance. Medical journal of Zambia. 2010;37(2):71-7.

53.	Kayigamba FR, Bakker MI, Lammers J, Mugisha V, Bagiruwigize E, Asiimwe A, et al. Provider-initiated HIV testing and counselling in Rwanda: acceptability among clinic attendees and workers, reasons for testing and predictors of testing. PloS one. 2014;9(4):e95459.

54.	Kennedy LA, Gordin FM, Kan VL. Assessing targeted screening and low rates of HIV testing. American journal of public health. 2010;100(9):1765-8.

55.	Khawcharoenporn T, Apisarnthanarak A, Phanuphak N. Active targeted HIV testing and linkage to care among men who have sex with men attending a gay sauna in Thailand. AIDS care. 2017;29(3):355-64.

56.	Kinsler JJ, Sayles JN, Cunningham WE, Mahajan A. Preference for physician vs. nurse-initiated opt-out screening on HIV test acceptance. AIDS care. 2013;25(11):1442-5.

57.	Kurth AE, Severynen A, Spielberg F. Addressing unmet need for HIV testing in emergency care settings: a role for computer-facilitated rapid HIV testing? AIDS education and prevention. 2013;25(4):287-301.

58.	Lander M, Tohani A, Dias A, O'Connell R. Assessing HIV testing in hepatitis: an audit of HIV testing uptake in a specialist hepatology clinic in an area of high prevalence for hepatitis B and C: P251. Hiv Medicine. 2014;15:96-7.

59.	Leblanc J, Hejblum G, Costagliola D, Durand-Zaleski I, Lert F, de Truchis P, et al. Targeted HIV screening in eight emergency departments: the DICI-VIH cluster-randomized two-period crossover trial. Annals of emergency medicine. 2018;72(1):41-53. e9.

60.	Leonard L, Berndtson K, Matson P, Philbin M, Arrington-Sanders R, Ellen JM. How physicians test: clinical practice guidelines and HIV screening practices with adolescent patients. AIDS Education and Prevention. 2010;22(6):538-45.

61.	Levin M, Mathema H, Stinson K, Jennings K. Acceptability, feasibility and impact of routine screening to detect undiagnosed HIV infection in 17-24-month-old children in the western sub-district of Cape Town. South African Medical Journal. 2012;102(4):245-8.

62.	Lolekha R, Kullerk N, Wolfe MI, Klumthanom K, Singhagowin T, Pattanasin S, et al. Assessment of a couples HIV counseling and testing program for pregnant women and their partners in antenatal care (ANC) in 7 provinces, Thailand. BMC international health and human rights. 2014;14(1):1-10.

63.	Lubelchek RJ, Kroc KA, Levine DL, Beavis KG, Roberts RR. Routine, rapid HIV testing of medicine service admissions in the emergency department. Annals of emergency medicine. 2011;58(1):S65-S70.

64.	Lucas KD, Eckert V, Behrends CN, Wheeler C, MacGowan RJ, Mohle-Boetani JC. Evaluation of routine HIV opt-out screening and continuum of care services following entry into eight prison reception centers—California, 2012. Morbidity and Mortality Weekly report. 2016;65(7):178-81.

65.	Luiken G, Joore I, Taselaar A, Schuit S, Geerlings S, Govers A, et al. Non-targeted HIV screening in emergency departments in the Netherlands. Neth J Med. 2017;75(9):386-93.

66.	Lyons MS, Lindsell CJ, Ledyard HK, Frame PT, Trott AT. Emergency department HIV testing and counseling: an ongoing experience in a low-prevalence area. Annals of emergency medicine. 2005;46(1):22-8.

67.	Lyss SB, Branson BM, Kroc KA, Couture EF, Newman DR, Weinstein RA. Detecting unsuspected HIV infection with a rapid whole-blood HIV test in an urban emergency department. JAIDS Journal of Acquired Immune Deficiency Syndromes. 2007;44(4):435-42.

68.	Malaju MT, Alene GD. Assessment of utilization of provider-initiated HIV testing and counseling as an intervention for prevention of mother to child transmission of HIV and associated factors among pregnant women in Gondar town, North West Ethiopia. BMC public health. 2012;12(1):1-8.

69.	Manjezi N, Fatti G, Mothibi E, Shaikh N, Oyebanji O, Grimwood A. WEAE0104 [LINE SEPARATOR] Index client trailing: a home‐based HIV counselling and testing strategy to identify and link people living with HIV to treatment. Journal of the International Aids Society. 2016;19.

70.	Mehta SD, Hall J, Lyss SB, Skolnik PR, Pealer LN, Kharasch S. Adult and pediatric emergency department sexually transmitted disease and HIV screening: programmatic overview and outcomes. Academic emergency medicine. 2007;14(3):250-8.

71.	Menacho I, Sequeira E, Muns M, Barba O, Leal L, Clusa T, et al. Comparison of two HIV testing strategies in primary care centres: indicator‐condition‐guided testing vs. testing of those with non‐indicator conditions. HIV medicine. 2013;14:33-7.

72.	Montoy J, Kaplan B, Dow W. 225 Can a Universal HIV Screening Policy Accomplish Targeted Testing? Results from a Trial of Monetary Incentives. Annals of Emergency Medicine. 2012;60(4):S82.

73.	Mwembo-Tambwe A, Kalenga M, Donnen P, Humblet P, Chenge M, Dramaix M, et al. HIV testing among women in delivery rooms in Lubumbashi, Democratic Republic of the Congo: a catch-up strategy for prevention of mother-to-child transmission. Revue d'epidemiologie et de sante publique. 2013;61(1):21-7.

74.	Myers JJ, Modica C, Dufour M-SK, Bernstein C, McNamara K. Routine rapid HIV screening in six community health centers serving populations at risk. Journal of general internal medicine. 2009;24(12):1269.

75.	Nakigudde R, Kabunga G, Sekyondwa M, Kawuma E, Karamagi Y, Odiit M, et al. Targeted HIV testing of children in the care of HIV positive adults, a gateway to the HIV positive child: The Mildmay, Uganda experience. International Journal of Infectious Diseases. 2014;21:406.

76.	Naugle DA, Dosso A, Tibbels NJ, Van Lith LM, Hendrickson ZM, Kouadio AM, et al. Addressing Uptake of HIV Testing and Linkage to Care Among Men in Côte d'Ivoire: An Evaluation of the Brothers for Life Program Implementation. JAIDS Journal of Acquired Immune Deficiency Syndromes. 2020;84(5):480-7.

77.	Nelwan EJ, Isa A, Alisjahbana B, Triani N, Djamaris I, Djaja I, et al. Routine or targeted HIV screening of Indonesian prisoners. International journal of prisoner health. 2016.

78.	Nsirim R, Ugochukwu G, Onuoha M, Okoroezi I, Ani C, Peters E. Effectiveness of provider-initiated testing and counseling in increasing HIV testing and counselling utilization and HIV detection rates in Ebonyi State, South-Eastern Nigeria. International journal of STD & AIDS. 2018;29(14):1362-7.

79.	Nunn A, Towey C, Chan PA, Parker S, Nichols E, Oleskey P, et al. Routine HIV screening in an urban community health center: results from a geographically focused implementation science program. Public Health Reports. 2016;131(1_suppl):30-40.

80.	Ogbo FA, Mogaji A, Ogeleka P, Agho KE, Idoko J, Tule TZ, et al. Assessment of provider-initiated HIV screening in Nigeria with sub-Saharan African comparison. BMC health services research. 2017;17(1):1-8.

81.	Osorio G, Hoenigl M, Quartarolo J, Barger K, Morris SR, Reed SL, et al. Evaluation of opt-out inpatient HIV screening at an urban teaching hospital. AIDS care. 2017;29(8):1014-8.

82.	Palfreeman A, Nyatsanza F, Farn H, McKinnon G, Schober P, McNally P. HIV testing for acute medical admissions: evaluation of a pilot study in Leicester, England. Sexually transmitted infections. 2013;89(4):308-10.

83.	Pai NP, Daher J, Prashanth H, Shetty A, Sahni RD, Kannangai R, et al. Will an innovative connected AideSmart! app-based multiplex, point-of-care screening strategy for HIV and related coinfections affect timely quality antenatal screening of rural Indian women? Results from a cross-sectional study in India. Sexually transmitted infections. 2019;95(2):133-9.

84.	Pant Pai N, Behlim T, Abrahams L, Vadnais C, Shivkumar S, Pillay S, et al. Will an unsupervised self-testing strategy for HIV work in health care workers of South Africa? A cross sectional pilot feasibility study. PloS one. 2013;8(11):e79772.

85.	Beasley D, Pontiff K, Bolton M, editors. Program Evaluation of Routine HIV Screening in the Emergency Department. Open Forum Infectious Diseases; 2015: Infectious Diseases Society of America.

86.	Prekker ME, Gary BM, Patel R, Olives T, Driver B, Dunlop SJ, et al. A comparison of routine, opt-out HIV screening with the expected yield from physician-directed HIV testing in the ED. The American journal of emergency medicine. 2015;33(4):506-11.

87.	Prekker M, Gary B, Olives T, Patel R, Gordon S, Schut R, et al. Non-targeted Opt-out Screening Using Rapid Tests for Human Immunodeficiency Virus in the Emergency Department: A Comparison With Physician-directed Testing: 299. Academic Emergency Medicine. 2011;18.

88.	Raman L, Duschl J, Wallis E, Orkin C. Effectiveness of opt-out HIV testing on a medical assessment unit in a high-prevalence area: P118. Hiv Medicine. 2015;16.

89.	da Costa Ribeiro LV, Sabidó M, Galbán E, de Oliveira Guerra JA, Mabey D, Peeling RW, et al. Home-based counseling and testing for HIV and syphilis–an evaluation of acceptability and quality control, in remote Amazonas State, Brazil. Sexually transmitted infections. 2015;91(2):94-6.

90.	Rodriguez V, Lester D, Connelly-Flores A, Barsanti FA, Hernandez P. Integrating routine HIV screening in the New York City community health center collaborative. Public Health Reports. 2016;131(1_suppl):11-20.

91.	Rosen DL, Wohl DA, Golin CE, Rigdon J, May J, White BL, et al. Comparing HIV case detection in prison during opt-in vs. opt-out testing policies. Journal of acquired immune deficiency syndromes (1999). 2016;71(3):e85.

92.	Russell S, Vernon S, Carson A, Harris D, Wheeler H. P127 HIV testing: are the targets off target? : BMJ Publishing Group Ltd; 2016.

93.	Russo G, Vita S, Miglietta A, Terrazzini N, Sannella A, Vullo V. Health profile and disease determinants among asylum seekers: a cross-sectional retrospective study from an Italian reception centre. Journal of Public Health. 2016;38(2):212-22.

94.	Sankoff J, Caruso E, Bender B, Haukoos J, Denver E. 216: Acceptance of Free Routine Opt-Out Rapid HIV Screening In the Emergency Department: Assessment of Race/Ethnicity and Payer Status. Annals of Emergency Medicine. 2010;56(3):S71.

95.	Sattin RW, Wilde JA, Freeman AE, Miller KM, Dias JK. Rapid HIV testing in a southeastern emergency department serving a semiurban-semirural adolescent and adult population. Annals of emergency medicine. 2011;58(1):S60-S4.

96.	Sau MS, Balamane M, Lurie M, Harwell J, Welle E, Mean C, et al. Assessment of prevention of mother-to-child transmission HIV services in the Bantey Meanchey Province in Cambodia. Journal of the International Association of Providers of AIDS Care (JIAPAC). 2016;15(4):345-9.

97.	Schlesinger S, Arora S, Menchine M, Newton K, Jacobson K, Takayama C, et al. 64 Routine, Non-Targeted Screening for HIV: Year One Findings of the “R/O HIV in the LAC+ USC ED” Program. Annals of Emergency Medicine. 2012;60(4):S24-S5.

98.	Schnall R, Liu N, Sperling J, Green R, Clark S, Vawdrey D, editors. Impact of an Electronic Alert for Non-targeted HIV Screening in the Emergency Department. NURSING RESEARCH; 2014: LIPPINCOTT WILLIAMS & WILKINS 530 WALNUT ST, PHILADELPHIA, PA 19106-3621 USA.

99.	Shih H-I, Ko N-Y, Hsu H-C, Wu C-H, Huang C-Y, Lee H-H, et al. Rapid Human Immunodeficiency Virus Screening in an Emergency Department in a Low HIV Seroprevalence Region. Japanese journal of infectious diseases. 2015:JJID. 2014.088.

100.	Silva A, Glick NR, Lyss SB, Hutchinson AB, Gift TL, Pealer LN, et al. Implementing an HIV and sexually transmitted disease screening program in an emergency department. Annals of emergency medicine. 2007;49(5):564-72.

101.	Control CfD, Prevention. Routine HIV screening during intake medical evaluation at a county Jail-Fulton County, Georgia, 2011-2012. MMWR Morbidity and mortality weekly report. 2013;62(24):495-7.

102.	Takano M, Iwahashi K, Satoh I, Araki J, Kinami T, Ikushima Y, et al. Assessment of HIV prevalence among MSM in Tokyo using self-collected dried blood spots delivered through the postal service. BMC infectious diseases. 2018;18(1):1-7.

103.	Tan XQ, Goh W-P, Venkatachalam I, Goh D, Sridhar R, Chan HC, et al. Evaluation of a HIV voluntary opt-out screening program in a Singapore hospital. Plos one. 2015;10(1):e0116987.

104.	Tlamsa A, Zucker J, Cennimo D, Sugalski G, Swaminathan S, editors. An Evaluation of Human Immunodeficiency Virus (HIV) Screening After Implementation of an Electronic Medical Record (EMR) Reminder. Open Forum Infectious Diseases; 2016: Oxford University Press.

105.	Truong H-HM, Akama E, Guzé MA, Otieno F, Obunge D, Wandera E, et al. Implementation of a community-based hybrid HIV testing services program as a strategy to saturate testing coverage in western Kenya. Journal of acquired immune deficiency syndromes (1999). 2019;82(4):362.

106.	Valenti SE, Szpunar SM, Saravolatz LD, Johnson LB. Routine HIV testing in primary care clinics: a study evaluating patient and provider acceptance. Journal of the Association of Nurses in AIDS Care. 2012;23(1):87-91.

107.	Veloso VG, Bastos FI, Portela MC, Grinsztejn B, João EC, Pilotto JHdS, et al. HIV rapid testing as a key strategy for prevention of mother-to-child transmission in Brazil. Revista de saude publica. 2010;44:803-11.

108.	Vohra R, Egan D, Ahearn M, Wall E, Wiener D. Are High-Risk Patients More Likely to Say Yes to an Human Immunodeficiency Virus Test? An Evaluation of Emergency Department Patients in a Rapid Testing Program: 700. Academic Emergency Medicine. 2013;20.

109.	Walensky RP, Arbelaez C, Reichmann WM, Walls RM, Katz JN, Block BL, et al. Revising expectations from rapid HIV tests in the emergency department. Annals of internal medicine. 2008;149(3):153-60.

110.	Wang Q, Chan P-L, Newman LM, Dou L-X, Wang X-Y, Qiao Y-P, et al. Acceptability and feasibility of dual HIV and syphilis point-of-care testing for early detection of infection among pregnant women in China: a prospective study. BMJ open. 2018;8(10):e020717.

111.	Wang X, Tang Z, Wu Z, Nong Q, Li Y. Promoting oral HIV self‐testing via the internet among men who have sex with men in China: a feasibility assessment. HIV medicine. 2020;21(5):322-33.

112.	Wheatley MA, Copeland B, Shah B, Heilpern K, Del Rio C, Houry D. Efficacy of an emergency department-based HIV screening program in the Deep South. Journal of Urban Health. 2011;88(6):1015-9.

113.	White DA, Cheung PT, Scribner AN, Frazee BW. A comparison of HIV testing in the emergency department and urgent care. The Journal of emergency medicine. 2010;39(4):521-8.

114.	White DA, Scribner AN, Schulden JD, Branson BM, Heffelfinger JD. Results of a rapid HIV screening and diagnostic testing program in an urban emergency department. Annals of emergency medicine. 2009;54(1):56-64.

115.	White DA, Scribner AN, Vahidnia F, Dideum PJ, Gordon DM, Frazee BW, et al. HIV screening in an urban emergency department: comparison of screening using an opt-in versus an opt-out approach. Annals of emergency medicine. 2011;58(1):S89-S95.

116.	White DA, Todorovic T, Petti ML, Ellis KH, Anderson ES. A comparative effectiveness study of two nontargeted HIV and hepatitis C virus screening algorithms in an urban emergency department. Annals of emergency medicine. 2018;72(4):438-48.

117.	Wilbur L, Huffman G, Lofton S, Finnell JT. The use of a computer reminder system in an emergency department universal HIV screening program. Annals of emergency medicine. 2011;58(1):S71-S3. e1.

118.	Xia Y-H, Chen W, Tucker JD, Wang C, Ling L. HIV and hepatitis C virus test uptake at methadone clinics in Southern China: opportunities for expanding detection of bloodborne infections. BMC Public Health. 2013;13(1):1-8.

119.	Yumo HA, Kuaban C, Ajeh RA, Nji AM, Nash D, Kathryn A, et al. Active case finding: comparison of the acceptability, feasibility and effectiveness of targeted versus blanket provider-initiated-testing and counseling of HIV among children and adolescents in Cameroon. BMC pediatrics. 2018;18(1):1-9.

120.	Zhu W, Mumby K, Dankerlui D, Manteuffel J, Ham C, Huang Y-LA, et al. Evaluation of a rapid point-of-care HIV screening program in an emergency department setting in Detroit, Michigan. Journal of clinical virology: the official publication of the Pan American Society for Clinical Virology. 2018;106:11-2.

121.	Zimet G, Cox A, Cox D, Harezlak J, Mays R, Fife R, et al. Rates and predictors of HIV testing among women attending Urban Health Clinics. Journal of the International Association of Physicians in AIDS Care. 2010;9(1):54.

122.	Zimmerman C, Thomas S, Batey D, Hamlin C, Ross-Davis K, Cantor R, et al. HIV Testing in the Emergency Department: Pilot Testing and Qualitative Assessment of Patient Perceptions: 172. Academic Emergency Medicine. 2010;17(5).

123.	Berhie SH, Tsai S, Miller ES, Garcia P, Yee LM. 667: Evaluation of state-mandated third-trimester repeat HIV testing in a single large tertiary care center. American Journal of Obstetrics & Gynecology. 2020;222(1):S424.

124.	Domínguez‐Berjón MF, Pichiule‐Castañeda M, García‐Riolobos MC, Esteban‐Vasallo MD, Arenas‐González SM, Morán‐Arribas M, et al. A feasibility study for 3 strategies promoting HIV testing in primary health care in Madrid, Spain (ESTVIH project). Journal of evaluation in clinical practice. 2017;23(6):1408-14.

125.	Hankin A, Freiman H, Copeland B, Shah B. 186 A Comparison of Routine HIV Screening Strategies in a Large, Inner-City Emergency Department: Integrated versus Parallel Models. Annals of Emergency Medicine. 2014;64(4):S67.

126.	Haukoos JS, Hopkins E, Bender B, Al‐Tayyib A, Long J, Harvey J, et al. Use of kiosks and patient understanding of opt‐out and opt‐in consent for routine rapid human immunodeficiency virus screening in the emergency department. Academic Emergency Medicine. 2012;19(3):287-93.

127.	Haukoos JS, Hopkins E, Conroy AA, Silverman M, Byyny RL, Eisert S, et al. Routine opt-out rapid HIV screening and detection of HIV infection in emergency department patients. Jama. 2010;304(3):284-92.

128.	Haukoos JS, Hopkins E, Bender B, Sasson C, Al-Tayyib AA, Thrun MW, et al. Comparison of enhanced targeted rapid HIV screening using the Denver HIV risk score to nontargeted rapid HIV screening in the emergency department. Annals of emergency medicine. 2013;61(3):353-61.

129.	Haukoos JS, Campbell JD, Conroy AA, Hopkins E, Bucossi MM, Sasson C, et al. Programmatic cost evaluation of nontargeted opt-out rapid HIV screening in the emergency department. PLoS One. 2013;8(12):e81565.

130.	Hechter RC, Bider-Canfield Z, Towner W. Effect of an electronic alert on targeted HIV testing among high-risk populations. The Permanente Journal. 2018;22.

131.	Hoxhaj S, Davila JA, Modi P, Kachalia N, Malone K, Ruggerio MC, et al. Using nonrapid HIV technology for routine, opt-out HIV screening in a high-volume urban emergency department. Annals of emergency medicine. 2011;58(1):S79-S84.

132.	Mahajan AP, Kinsler JJ, Cunningham WE, James S, Makam L, Manchanda R, et al. Does the centers for disease control and prevention’s recommendation of opt-out HIV screening impact the effect of stigma on HIV test acceptance? AIDS and Behavior. 2016;20(1):107-14.

133.	Mbopi-Keou F-X, Kalla G, Guiadem R, Tchouamani H, Mbele R, Nkada C, et al. Large scale HIV survey in Cameroon by mass HIV testing mobile units: Evidence of HIV epidemic hot spot areas and high HIV vulnerability of women over time. International Journal of Infectious Diseases. 2010;14:e79-e80.

134.	McGuire R, Moore E. Using a configurable EMR and decision support tools to promote process integration for routine HIV screening in the emergency department. Journal of the American Medical Informatics Association. 2016;23(2):396-401.

135.	Schrantz SJ, Babcock CA, Theodosis C, Brown S, Mercer S, Pillow MT, et al. A targeted, conventional assay, emergency department HIV testing program integrated with existing clinical procedures. Annals of emergency medicine. 2011;58(1):S85-S8. e1.

136.	Yeganeh N, Simon M, Dillavou C, Varella I, Santos BR, Melo M, et al. HIV testing of male partners of pregnant women in Porto Alegre, Brazil: a potential strategy for reduction of HIV seroconversion during pregnancy. Aids Care. 2014;26(6):790-4.

137.	Chaffin M, Balachova T, Shaboltas A, Bohora S, Batluk J, Bonner B, et al. Increasing HIV testing as a prevention strategy: a randomized trial of opt-in and opt-out HIV testing strategies among at-risk women. Alcoholism: Clinical and Experimental Research. 2016;40(S1):60A.

138.	Chamie G, Ndyabakira A, Marson KG, Emperador DM, Kamya MR, Havlir DV, et al. A pilot randomized trial of incentive strategies to promote HIV retesting in rural Uganda. PloS one. 2020;15(5):e0233600.

139.	Chamie G, Schaffer E, Ndyabakira A, Emperador D, Kwarisiima D, Havlir DV, et al. A randomized trial of novel strategies to incentivize HIV testing among men in Uganda. Topics in Antiviral Medicine. 2017;25(1 Supplement 1):15s.

140.	Chesang K, Agot K, Kimani J, Muthumbi G, Gichangi P, Musyoki H, et al., editors. Using Peer Educators to Scale-up HIV Oral Self-testing among Female Sex Workers: An Implementation Science Approach from Kenya. AIDS RESEARCH AND HUMAN RETROVIRUSES; 2016: MARY ANN LIEBERT, INC 140 HUGUENOT STREET, 3RD FL, NEW ROCHELLE, NY 10801 USA.

141.	Cowan FM, Davey C, Fearon E, Mushati P, Dirawo J, Chabata S, et al. Targeted combination prevention to support female sex workers in Zimbabwe accessing and adhering to antiretrovirals for treatment and prevention of HIV (SAPPH-IRe): a cluster-randomised trial. The lancet HIV. 2018;5(8):e417-e26.

142.	Gillet C, Darling KE, Senn N, Cavassini M, Hugli O. Targeted versus non-targeted HIV testing offered via electronic questionnaire in a Swiss emergency department: A randomized controlled study. PloS one. 2018;13(3):e0190767.

143.	Haukoos J, Lyons M, Rothman R. The HIV Tested Trial: A Multi-center Pragmatic Randomized Comparison of HIV Screening Strategy Effectiveness inthe Emergency Department. Acad Emerg Med. 2017;24(S1):S42-3.

144.	Kelvin EA, George G, Mwai E, Nyaga EN, Mantell JE, Romo ML, et al. Offering self-administered oral HIV testing as a choice to truck drivers in Kenya: predictors of uptake and need for guidance while self-testing. AIDS and Behavior. 2018;22(2):580-92.

145.	Lyons MS, Lindsell CJ, Ruffner AH, Wayne DB, Hart KW, Sperling MI, et al. Randomized comparison of universal and targeted HIV screening in the emergency department. Journal of acquired immune deficiency syndromes (1999). 2013;64(3):315.

146.	Merchant RC, Clark MA, Langan IV TJ, Mayer KH, Seage III GR, DeGruttola VG. Can computer-based feedback improve emergency department patient uptake of rapid HIV screening? Annals of emergency medicine. 2011;58(1):S114-S9. e2.
